# Livestock, pathogens, vectors, and their environment: a causal inference-based approach to estimating the pathway-specific effect of livestock on human African trypanosomiasis risk

**DOI:** 10.1101/2022.01.13.22268994

**Authors:** Julianne Meisner, Agapitus Kato, Marshall Lemerani, Erick Mwamba Miaka, Acaga Ismail Taban, Jonathan Wakefield, Ali Rowhani-Rahbar, David Pigott, Jonathan Mayer, Peter Rabinowitz

## Abstract

Livestock are important reservoirs for many diseases, and investigation of such zoonoses has long been the focus of One Health research. However, the effects of livestock on human and environmental health extend well beyond direct disease transmission.

In this retrospective ecological cohort study we use pre-existing data and methods derived from causal inference and spatial epidemiology to estimate three hypothesized mechanisms by which livestock can come to bear on human African trypanosomiasis (HAT) risk: the reservoir effect, by which infected cattle and pigs are a source of infection to humans; the zooprophylactic effect, by which preference for livestock hosts exhibited by the tsetse fly vector of HAT means that their presence protects humans from infection; and the environmental change effect, by which livestock keeping activities modify the environment in such a way that habitat suitability for tsetse flies, and in turn human infection risk, is reduced. We conducted this study in four high burden countries: at the point level in Uganda, Malawi, and Democratic Republic of Congo (DRC), and at the county-level in South Sudan.

Our results indicate cattle and pigs play an important reservoir role for the rhodesiense form (rHAT) in Uganda, however zooprophylaxis outweighs this effect for rHAT in Malawi. For the gambiense form (gHAT) we found evidence that pigs may be a competent reservoir, however dominance of the reservoir versus zooprophylactic pathway for cattle varied across countries. We did not find compelling evidence of an environmental change effect.

**Author summary:** One Health research is most commonly interested in livestock as reservoirs of zoonotic diseases (i.e., infectious diseases transmissible from animals to humans), however livestock also exert environmental effects on a range of scales. At a local level, grazing and brush-clearing activities related to livestock keeping can reduce vegetation and increase temperature, in turn reducing habitat for and density of disease reservoirs and vectors. Furthermore, many arthropod vectors of human (zoonotic and non-zoonotic) diseases exhibit host species preference; when livestock hosts are preferred, the presence of these animals may reduce the risk of human infection. When all three of these effects act in concert, their relative strength governs whether the overall effect of livestock on human disease risk is positive (harmful) or negative (protective), a balance which is likely focus-specific.

Using pre-existing data and methods drawn from causal inference and spatial epidemiology, we estimate the contribution of these three pathways to the effect of cattle and pigs on human African trypanosomiasis (HAT) risk in Uganda, Malawi, Democratic Republic of Congo, and South Sudan. We find little evidence of an environmental change effect, however cattle and pigs appear to play both reservoir and zooprophylactic roles in the epidemiology of this disease.

## Introduction

Domestic and wild animals—in particular bovids such as cattle, bushbuck, and African buffalo, and suids such as domestic pigs and warthogs—are known to be important reservoirs for the acute form of human African trypanosomiasis, caused by *Trypanosoma brucei rhodesiense*(rHAT) [1–4], and there is some evidence that domestic pigs may also be reservoirs for the chronic form, caused by *T. b. gambiense* (gHAT).

However, livestock may also influence the distribution of HAT beyond their role as reservoir hosts. HAT is transmitted by the tsetse fly, and several species which are important HAT vectors have been documented to prefer animal hosts, and to only bite humans during times of stress [5]. Also appreciated with the malaria vector *Anopheles arabiensis* [6–8], this preference may shield humans from tsetse bites and thus HAT exposure when favored animal hosts are nearby, termed zoopropylaxis. The fly biting behaviors that favor a zooprophylactic role versus a reservoir role are distinct, with strongly zoophilic tsetse being unlikely to be competent HAT vectors, and animal reservoirs unlikely to be important where the predominant HAT vector is strongly anthrophilic. Furthermore, zoonotic transmission requires host switching—whereby a given fly feeds on a human host, followed by an animal host, then subsequently on a human host—while zooprophylaxis does not [9]. Tsetse preference for animal hosts is thus harmful (up to a threshold) for HAT control in the presence of animal reservoir, and helpful in its absence. Consistent with this, gHAT models have found that in the absence of animal reservoirs tsetse biting preference for animals had a negative effect on HAT risk, while in the presence of animal reservoirs biting preference for humans became more influential [9].

Livestock grazing can bring about ecosystem degradation, in the form of desertification (the conversion of fertile land to desert) in arid climates, increased woody plant cover in semi-arid rangelands, and deforestation in humid climates [10]. As early as 1988, it was noted that overgrazing could potentially change local and global weather systems [11]. Insect vectors of public health importance, including tsetse fly vectors of HAT, are sensitive to environmental factors [12]. In Burkina Faso and Ghana, cattle keeping, crop farming, and other climactic and anthropogenic factors have altered riverine landscapes, fragmenting gallery forests and influencing tsetse distribution [13]. Similarly, in Ethiopia, where animal African trypanosomiasis (AAT) is endemic but HAT is not, regulatory changes leading to expansion in agricultural land was documented to alter the distribution of tsetse flies [14]. Furthermore, the clearing of bush by subsistence livestock agriculture in pre-colonial Africa is credited with small-scale control of tsetse, termed “agricultural prophlyaxis,” and the resurgence of bush following catastrophic livestock losses to rinderpest (“cattle plague”) is credited with the sleeping sickness outbreaks of the early 20th century [11].

Livestock may thus come to bear on HAT distribution through three hypothesized pathways: disease reservoir, zooprophylaxis, and environmental change. While One Health approaches to HAT control target the reservoir pathway, the environmental change pathway was successfully employed for HAT and AAT control in pre-colonial Africa through agricultural prophylaxis, and in colonial Africa through environmentally-traumatic approaches such as brush clearing and widespread use of persistent organochlorine insecticides. Furthermore, vector control strategies that target livestock, such as insecticide treatment of cattle (ITC), exploit the close proximity of cattle to humans and their need to travel daily to the riverine habitats favored by gHAT vectors for water, and thus zooprophylaxis [15, 16].

Estimation of pathway-specific effects can narrow knowledge gaps surrounding the epidemiology of HAT, facilitate the targeted deployment of control measures, and provide key parameters for mathematical modeling. Previous authors have estimated elements of these effects in select foci, including the predictive ability of remote-sensed environmental data for tsetse distribution in Nuguruman, Kenya [17] and the effects of trypanocide mass-treatment of cattle on rHAT risk in Uganda [18], of habitat fragmentation on rHAT risk in Eastern Zambia [19], and of animal husbandry and climate on AAT risk in Burkina Faso [20]. However, we are aware of no study that seeks to disentangle the mechanisms driving the effect of livestock on HAT risk. Thus, significant uncertainty remains surrounding the role of pigs as reservoirs of gHAT and the ability of both pigs and cattle to bring about environmental changes that in turn affect the distribution of HAT.

In this study, we apply the mediational g-formula [21] to estimate the magnitude of the direct and indirect effects of cattle and pig density on HAT risk in three high-burden countries: Malawi (rHAT), Uganda (rHAT and gHAT), and Democratic Republic of Congo (DRC, gHAT), using remote-sensed data, HAT surveillance data collated by the WHO Atlas of HAT, and a new time-series of high resolution livestock density maps. We define livestock density as the ratio of livestock (separately for cattle and pigs) to humans, and fit all models separately for each country, for gHAT and rHAT, and for two mediators: normalized difference vegetation index (NDVI), a measure of vegetation cover, and land surface temperature (LST). Additionally, in South Sudan (gHAT) we use a spatial extension of the regression approach to mediation analyses described by VanderWeele [21], and a single county-level map of livestock density produced for 2008.

## Materials and methods

Our study is a retrospective ecological cohort study with cluster-year (0.017° pixel) as the unit of analysis in Malawi, Uganda, and DRC, and county (administrative level 2) as the unit of analysis in South Sudan. This distinction was necessitated by the absence of exposure (livestock ownership) data with cluster-level geolocation in South Sudan.

### Study population

In Uganda, DRC, and Malawi, our study population included all clusters within five hours’ travel time of a fixed health facility capable of HAT diagnosis [22]. Study areas were defined separately for gHAT and rHAT in Uganda. In South Sudan all counties which reported one or more HAT cases or conducted active surveillance during the study period comprised the study area.

The study period was defined separately for each country, on the basis of WHO Atlas of HAT data access provided to the authors and available exposure data: 2006-2018 for Uganda, 2003-2014 for Malawi, and 2010-2013 for DRC. In South Sudan the study period was restricted to 2008 alone as only the 2008 census provided geolocated (county-level) livestock ownership data for this country (detailed in S1 File).

### DAG formulation

In all study countries we adopted the counterfactual approach to mediation analysis, identifying confounders by the DAG depicted in Fig 1. Index events are natural disasters or armed conflicts, and proximity to protected areas—a proxy for density of wildlife reservoirs of HAT—refers to rHAT models only. Time-varying variables are indexed by *t*, and variables in the minimally-sufficient adjustment set are boxed: {elevation, index events, LST, NDVI, wealth, vector control}.

**Fig 1.**
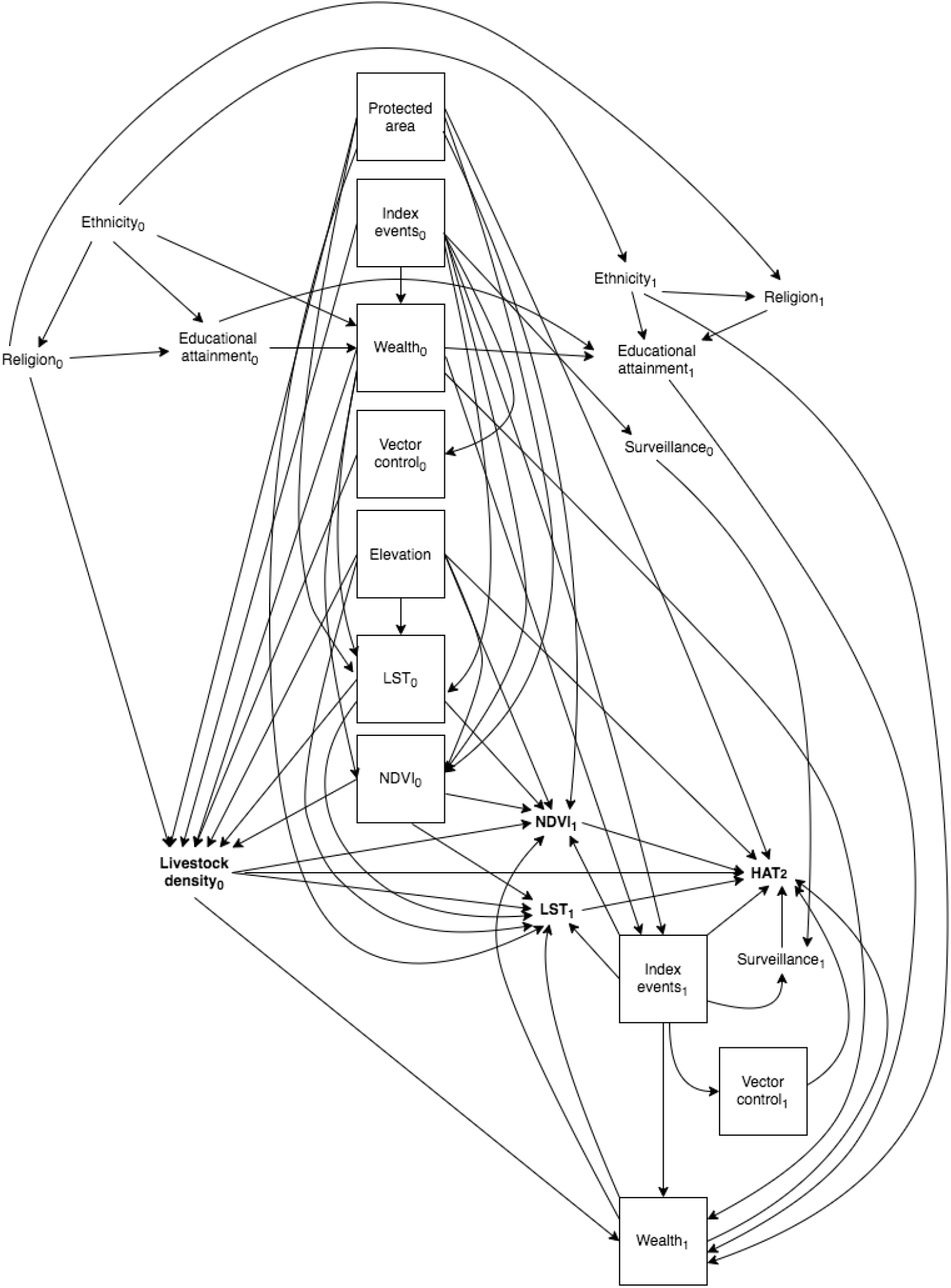
Final time-varying directed acyclic graph (DAG), restricted to two time points. Index events are natural disasters or armed conflicts. Protected areas pertains to rHAT models only. Exposure and outcome of interest are bolded, and confounders in the minimally-sufficient set are denoted by solid boxes.

Our DAG encodes our beliefs about the underlying temporal structure of these causal relationships. LST, NDVI, and wealth exhibit exposure-confounder feedback, where these variables at time *t* are a cause of livestock density at time *t*, which in turn is a cause of LST, NDVI and wealth at time *t* + 1. In the presence of exposure-confounder feedback confounding cannot be controlled using adjustment in regression models. Here we apply the mediational g-formula, one of Robin’s generalized methods (“g-methods”), which allows estimation of causal effects in the face of time-varying confounding with exposure-confounder feedback [23–25]. As we did not have longitudinal data in South Sudan this issue did not arise; in this country, we applied the regression approach to mediation [23].

Note that while ethnicity, educational attainment, and religion are generally not time-varying at an individual-level, in our application these represent group-level variables (i.e., proportion receiving higher than a given level of education, proportion belonging to the majority ethnic group, and proportion belonging to Muslim, Christian, and other religions) and thus do vary in time.

### Measures

#### Exposure

Exposure is livestock density, parameterized as the ratio of livestock (cattle and pigs, separately) to humans using maps we detail in a separate publication (S1 File). In Malawi, Uganda, and DRC these were a time-series of continuous (raster) maps. Due to limited data availability, in South Sudan we produced a single areal (county-level) map for 2008.

#### Outcome data

Outcome data included all new cases of HAT diagnosed in a given year and given cluster (Malawi, Uganda, DRC) or county (South Sudan) in the WHO Atlas of HAT [26]. Clusters which did not appear in the Atlas were either assigned 0 cases if they were at least 1km from any clusters with reported cases, and removed from all analyses otherwise. Denominator data came from the University of Southhampton’s WorldPop project [27].

#### Confounders

We estimated wealth using an exploratory factor analysis approach modeled after the DHS Wealth Index [28] but excluding livestock-related measures. We then used spatial modeling to generate continuous annual maps in Uganda, Malawi, and DRC, and a single areal map for 2008 for South Sudan (detailed in S1 Appendix).

Natural disasters and armed conflicts were identified using the EM-DAT and UCDP/PIRO Armed Conflict Databases, respectfully [29, 30]. NDVI data came from the Land Long Term Data Record 5 (LTDR5) [31]; LST data came from MODIS/Terra MOD21 for 2000-2002 [32], and from MODIS/Aqua MYD21 for 2003-2014 [33]. Finally, elevation data came from GMTED2010 [34], with the median product used.

We were not able to adjust for vector control as top-down vector control efforts remain limited [35, 36] and it is not practical to parameterize farmer-led efforts on the scale of this study.

### Causal estimands

In keeping with the potential outcomes framework for causal inference, we define estimands in terms of counterfactual outcomes 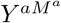, where *Y* denotes outcome, *M* denotes mediator, and *A* = *a* denotes exposure fixed at level *a*. Under the identifiability criteria, population-average causal estimands can be derived from observational data.

The parameters of interest are the natural direct and natural indirect effects each for LST and NDVI. The natural direct effect is the difference in counterfactual outcomes with the mediator held fixed at the value (*M*^*a*\*^) it would take under exposure value *A* = *a* (for a binary exposure, this is nominally *A* = 0, or unexposed), and the exposure varied from *A* = *a \** to *A* = *a*. In counterfactual notation, the population average natural direct effect is expressed as 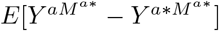. The natural indirect effect is the difference in counterfactual outcomes with the exposure fixed at *A* = *a*, but the mediator varied from the value it would take under exposure *A* = *a* vs.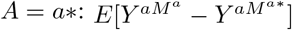. These effects are not identifiable when there is exposure-confounder feedback, however their randomized interventional analogs, which compare counterfactual outcomes under mediator values derived from random draws of the mediator distribution under exposure *A* = *a* vs. *A* = *a*\* (*G*^*a*^ and *G*^*a*\*^, respectively), can be identified [21]. The resultant natural direct effect analog is 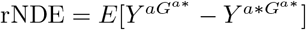, and the natural indirect effect analog is 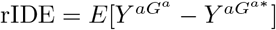. For notational simplicity, we will continue to use the 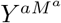 notation. We defined *A* = *a*\* as mean livestock density, and *A* = *a* as mean × 1.5, and specify our causal estimand on the ratio scale (detailed in S2 Appendix), generating a rate ratio analog corresponding to a 50% increase in exposure.

### Mediational g-formula

The mediational g-formula uses model-based standardization to derive these estimands; its implementation, along with the identifiability criteria, are detailed in S2 Appendix. Briefly, we fit a series of `glm()` models in R for each time-varying confounder (including the two mediators in question) and for outcome, separately for each year and ensuring control is only for the past. We then simulated from each model in turn using the `predict()` function and the level of exposure (livestock density) specified by the causal estimand in question. For instance, as the natural direct effect requires estimation of 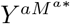, we fixed exposure at *A* = *a* for the outcome and confounder models, and at *A* = *a*\* for the mediator models. We then tied these models together using the law of total probability, marginalized to obtain the causal parameter 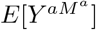, and produced uncertainty bounds through non-parametric bootstrapping (100 simulations). Note that implementation was conducted separately for each mediator.

As the no interference assumption is likely violated in our study, we instead assume partial interference, in which each unit’s potential outcome is independent of the rest conditional on a specified interference set *χ*_*i*_ = {*i*_1_, *i*_2_, …} [37, 38]. Reflecting the range of tsetse flies and grazing ranges of livestock, we define the interference set as all other clusters within a 5km radius [39, 40].

Our targets of inference are:

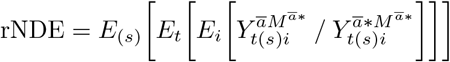

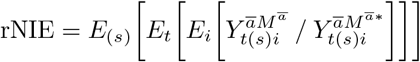

where *s* indexes simulation (100 in total), t indexes year, *i* indexes cluster, and 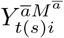 is the one-year cumulative incidence of HAT for a given livestock species, under *A* = *a*.

Validation of the mediational g-formula is conducted through implementation of the “natural course” model, whereby exposure (livestock density) is modeled as for the time-varying confounders. If the observed distribution is recovered, model performance is deemed adequate.

### Regression approach

Due to the absence of longitudinal exposure data in South Sudan, we applied a regression approach to our mediation analysis in this country. Under this approach, two models are fit—one for outcome, and one for mediator—and causal estimands are derived from linear combinations of parameters from these models.

Consistent with our DAG formulation, we assume that wealth measured in the 2008 South Sudan census is upstream of livestock density ascertained in the same census. Thus, wealth at time *t*_1_ constitutes a mediator-outcome confounder which is downstream of exposure (as livestock density is a cause of wealth), precluding a simple regression approach to mediation analysis [23]. However, wealth was only measured at time 0 (2008), thus we proceeded with the regression approach and acknowledge our results will not have a causal interpretation.

The outcome (Poisson) model was as follows, where *c* indexes county:

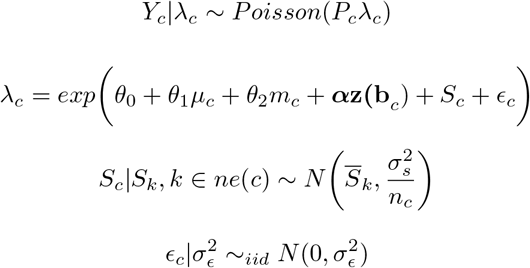

where

- *Y*_*c*_ is the number of cases in county *c* in 2012
- *m*_*c*_ is the mediator value in county *c* in 2010, *M* ∈{NDVI, LST}
- *µ*_*c*_ is estimated livestock density in county *c* in 2008
- ***α*** is a vector of coefficients
- **z(b***c*) is a vector of exposure-outcome, exposure-mediator, and mediator-outcome confounders, *k* ∈ {2008 wealth; 2007 NDVI, LST, conflicts, disasters; 2009 conflicts, disasters}
- *S*_*c*_ are county-level structured random effects which follow the ICAR model with marginal variance 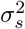
- *ne*(*c*) denotes neighbors (shared boundary) of county *c*
- *n*_*c*_ is the number of neighbors of county *c*
- *∈*_*c*_ are unstructured (iid) county-level spatial random effects
- *P*_*c*_ is the offset, given as population in county *c*

and the mediator (linear) model was:

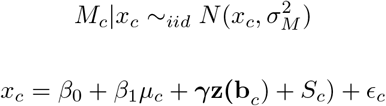

where

- *M*_*c*_ is the mediator value in county *c, M* ∈{2010 NDVI, 2010 LST}
- *µ*_*c*_ is estimated livestock density in county *c* in 2008
- ***γ*** is a vector of coefficients
- **z(b***c*) is a vector of exposure-mediator confounders, *k* ∈ {2008 wealth; 2007 NDVI, LST, conflicts, disasters}
- *S*_*c*_ are structured county-level spatial random effects, assumed to follow the ICAR model
- *∈*_*c*_ are unstructured (iid) county-level spatial random effects

and ICAR is the intrinsic conditional autoregressive model, which smooths each county’s random effect to that of its neighbors.

These models do not contain an exposure × mediator interaction term (imposing the assumption of linearity in the direct and indirect effects of exposure on outcome across levels of the mediator), thus the direct effect is then calculated as:

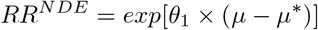

and the indirect effect is calculated as:

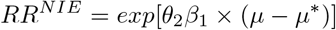

where *µ* − *µ*\* are the two values of livestock density we are comparing [23]. We have parameterized exposure such that this contrast corresponds to a 50% increase in livestock density, allowing direct comparison with mediational g-formula results in the other countries. We used penalized complexity (PC) priors for the smoothing model, with *Pr*(*σ*_*∈*_) *>* 1 = *Pr*(*σ*_*s*_) *>* 1 = 0.01. This yields a posterior 99% credible interval for each random effect’s residual rate ratio of (0.36, 2.71) [41, 42].

We first fit naive models which did not account for measurement error in livestock density or wealth arising from the fact that these variables were estimated. We subsequently re-fit these specifying livestock density and wealth as random effects following a classical measurement error (MEC) model. We detail implementation of this model in S3 Appendix.

No off-the-shelf software is available to calculate standard errors for these estimands in the presence of spatial structure, thus we derived uncertainty estimates by taking 1,000 draws from the posterior distribution of each parameter in the above models, and taking the 2.5% and 97.5% percentiles of this distribution as the posterior 95% credible interval.

### Data and code availability

HAT outcome data can be requested from the WHO (https://www.who.int/trypanosomiasis_african/country/foci_AFRO/en/) and livestock density maps can be downloaded from https://github.com/JulianneMeisnerUW/LivestockMaps. Mediator and confounder data are available for download from the websites linked in our References. All analyses were performed in R, and all code are available in the same GitHub repository.

## Results

### Descriptive statistics

After removing clusters which did not meet the inclusion criteria, we were left with 3,982 clusters/year in Malawi, 5,746 clusters/year for gHAT models in Uganda, 13,570 clusters/year for rHAT models in Uganda, and 247,205 clusters/year in DRC. In South Sudan, the study area was comprised of 17 counties. General study areas are presented in Fig 2.

**Fig 2.**
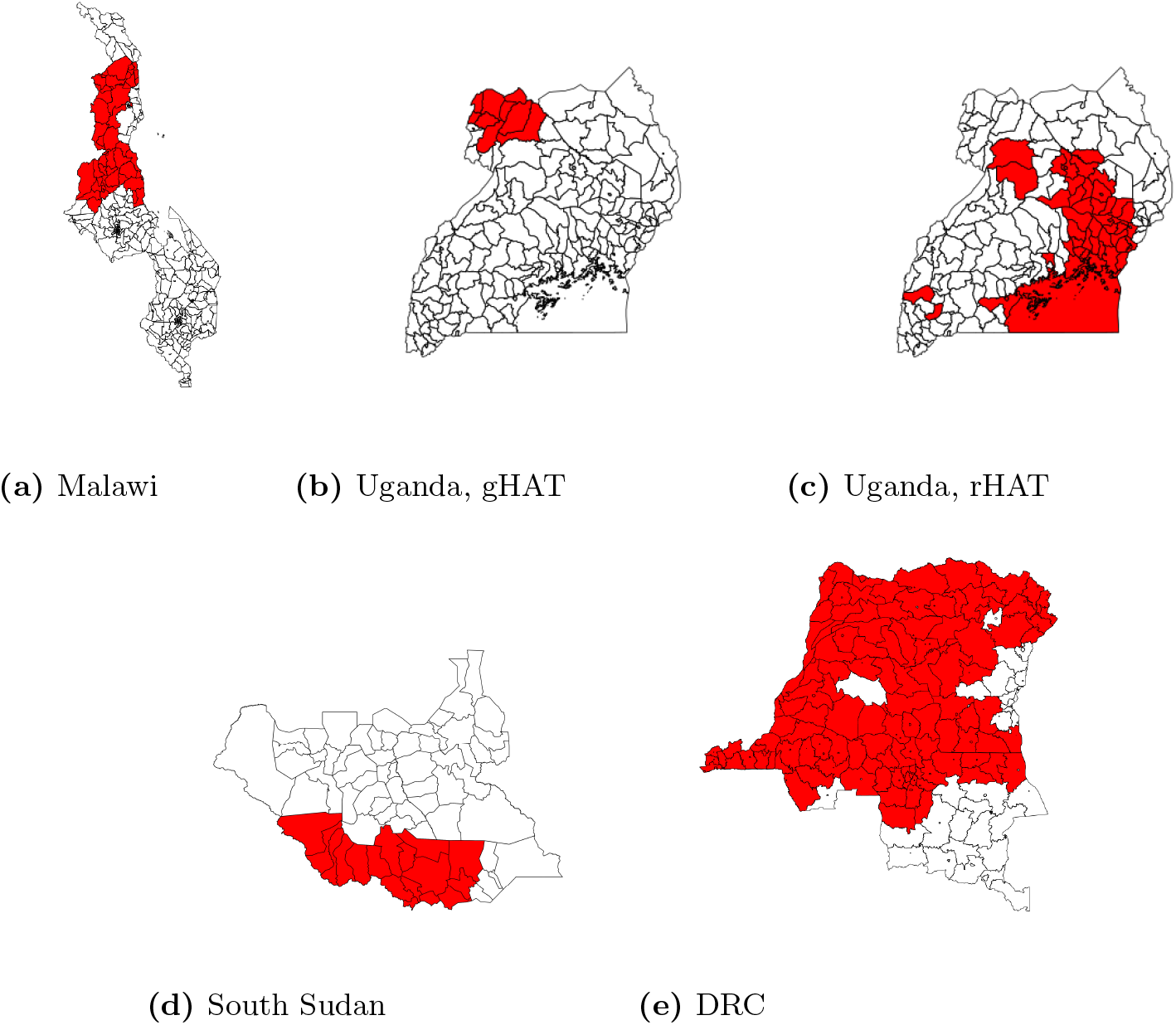
National maps with administrative areas represented in the study data highlighted in red. (a) Malawi: traditional authority (administrative level 3); (b-d) Uganda and South Sudan: county (administrative level 2); (e) DRC: territory (administrative level 2)

Malawi did not have any armed conflicts during the study period, and South Sudan did not have any naturaldisasters. Descriptive statistics are presented in Tables 1 and 2, with additional descriptive statistics figures presented in S4 Appendix.

**Table 1.** Descriptive statistics, Malawi, Uganda, and DRC.

| Variable | Mean (sd) |  |  |  |
| --- | --- | --- | --- | --- |
|  | Malawi | Uganda, gHAT | Uganda, rHAT | DRC |
| Cattle density | 0.09 (0.31) | 0.31 (0.92) | 0.16 (0.42) | 0.06 (0.06) |
| Pig density | 0.09 (0.19) | 0.24 (0.72) | 0.06 (0.10) | 0.06 (0.03) |
| Elevation (meters) | 1,125 (280) | 890 (166) | 1,099 (79) | 524 (194) |
| HAT cases | 0.01 (0.14) | 0.04 (0.35) | 0.01 (0.15) | 0.02 (0.35) |
| LST (K) | 307 (4.7) | 281 (96) | 277 (95) | 306 (3.67) |
| NDVI | 0.15 (0.14) | 0.28 (0.13) | 0.26 (0.21) | 0.22 (0.18) |
| Wealth | 1.30 (1.04) | 0.84 (0.03) | 0.85 (0.03) | 0.25 (0.22) |
| Conflict | 0 (0%)* | 743 (68%)* | 1,824 (71%)* | 177,468 (9%)* |
| Disaster |  |  |  |  |
| Flood | 19,548 (33%)* | 413 (38%)* | 2,290 (0.9%)* | 46,921 (2.37%)* |
| Storm | 1,213 (2%)* | 0 (0%)* | 0 (0%)* | 0 (0%)* |
| Epidemic | 0 (0%)* | 2,065 (1.9%)* | 6,307 (2.5%)* | 28,343 (1.43%)* |
| Landslide | 0 (0%)* | 0 (0%)* | 127 (0.05%)* | 0 (0%)* |
| Drought | 0 (0%)* | 0 (0%)* | 8 (<0.01%)* | 0 (0%)* |
| Earthquake | 0 (0%)* | 0 (0%)* | 0 (0%)* | 410 (0.02%)* |
| Wildfire | 0 (0%)* | 0 (0%)* | 0 (0%)* | 16,575 (0.84%)* |

Descriptive statistics over study clusters and period (2000-2014 for Malawi, 2000-2018 for Uganda and DRC). sd: standard deviation. \*n(%)

**Table 2.** Descriptive statistics, South Sudan.

| Variable | Mean (sd) |
| --- | --- |
| Cattle density | 0.77 (1.15*) |
| Pig density | 0.02 (1.25*) |
| Elevation (meters) | 693 (158) |
| HAT cases | 34 (52) |
| Number screened | 2,953 (4,200) |
| LST (K) | 312.54 (3.22) |
| NDVI | 0.28 (0.07) |
| Wealth | 0.19 (0.004*) |
| Conflict | 8 (47%)** |

Descriptive statistics over study counties. 2008 data: cattle density, pig density, HAT cases, number screened, WorldPop population, LandScan population, wealth. 2007 data: LST, NDVI. 2006 data: conflicts. sd: standard deviation. \*Mean of design-based standard error.\*\*n, (%)

### Mediational g-formula

Validation results indicated very strong agreement between the natural course model and reported HAT cases with the exception of 2010 and 2012 for the pig-gHAT models in Uganda. We removed these years from the Uganda analyses; across the remaining years and models, the mean of the squared difference between observed livestock density and that predicted by the “natural course” model across cluster-years was *<* 1*x*10^−4^.

Results are presented in Tables 3 - 5. In Uganda, we found no evidence of an indirect effect across both mediators, livestock species, and HAT types. Direct effects in Uganda were positive for pigs in both gHAT and rHAT foci, positive for cattle in rHAT foci, and negative for cattle in gHAT foci, however these effects only reached statistical significance for the pig-rHAT effect. DRC results were similar to the Uganda gHAT results (i.e., no evidence of an indirect effect, positive direct effects for pigs), however we also detected a positive direct effect for cattle—albeit weaker than for pigs—and all direct effects reached statistical significance. In Malawi, we did not run LST models due to large amounts of missingness, and we removed location within a protected area, elevation, and disaster from all models as very few HAT cases were observed during the study period leading to concerns regarding overfitting. For NDVI in this country, indirect effects were close to null and non-significant; direct effects were positive for both cattle and pigs, being stronger and statistically significant for pigs.

**Table 3.**
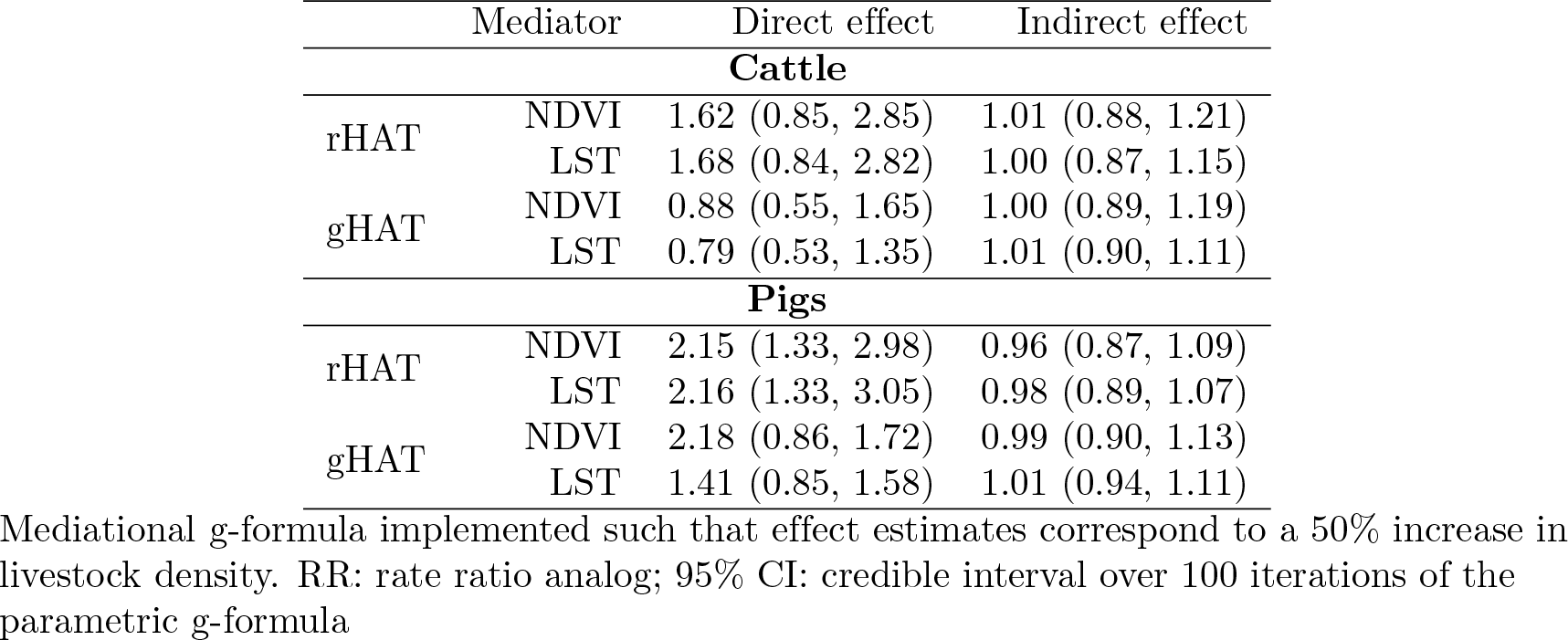
Mediation analysis results, Uganda RR (95% CI)

**Table 4.**
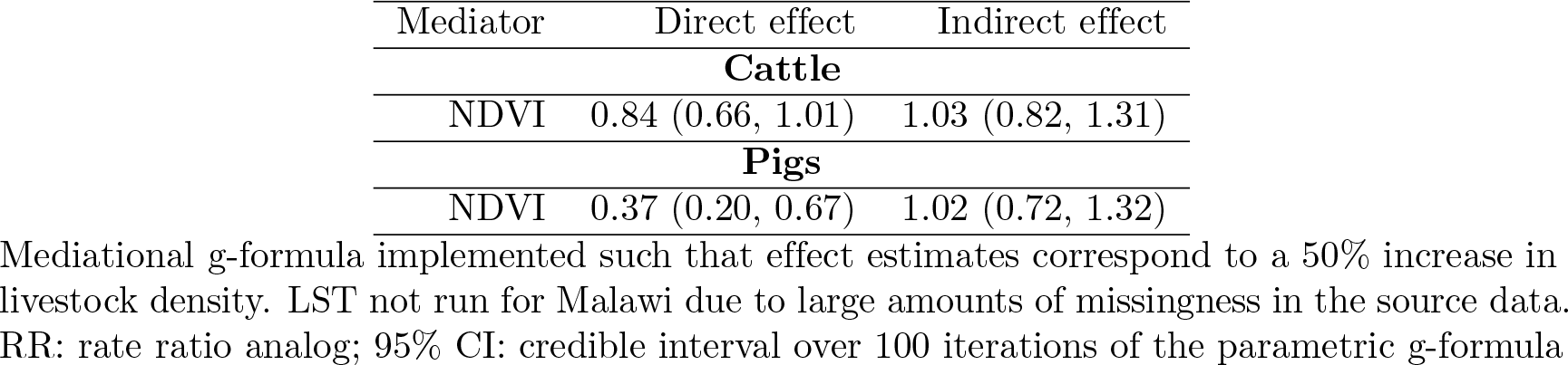
Mediation analysis results, Malawi RR (95% CI)

**Table 5.**
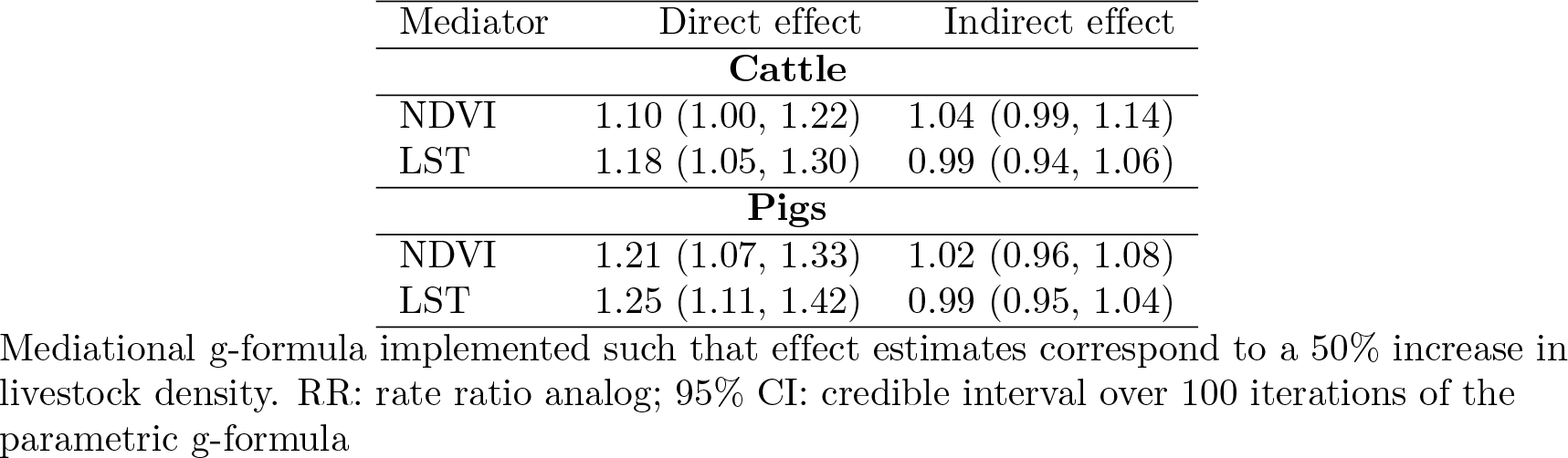
Mediation analysis results, DRC RR (95% CI)

### Regression

As the main models are expected to be biased, we will only interpret the MEC models here. Indirect effects were strong and negative for pigs for both mediators and for cattle for LST only; direct effects were strong and negative for cattle, but weaker and on both sides of the null for pigs. However in all cases uncertainty bounds were very wide and no effects reached statistical significance. Results are presented in Table 6.

**Table 6.** Mediation analysis results, South Sudan RR (95% CI)

| Mediator | Model | Direct effect | Indirect effect |
| --- | --- | --- | --- |
| Cattle |  |  |  |
| NDVI | Naive | 0.34 (0.09, 1.28) | 0.99 (0.46, 1.60) |
| LST | Naive | 0.32 (0.12, 0.80) | 0.73 (0.09, 2.88) |
| NDVI | MEC | 0.22 (0.13, 0.41) | 0.98 (0.38, 1.55) |
| LST | MEC | 0.28 (0.17, 0.45) | 0.50 (0.09, 1.54) |
| Pigs |  |  |  |
| NDVI | Naive | 0.96 (0.09, 8.85) | 0.75 (0.13, 3.59) |
| LST | Naive | 0.93 (0.26, 3.05) | 0.81 (0.13, 3.50) |
| NDVI | MEC | 1.55 (0.52, 3.68) | 0.62 (0.09, 1.35) |
| LST | MEC | 0.86 (0.44, 1.68) | 0.57 (0.13, 2.15) |

Estimates of direct (natural or controlled) effect and natural indirect effect, using 2012 HAT data as outcome, 2008 livestock density data as exposure, and 2010 NDVI and LST data as mediators, fit in two separate models. Adjustment performed for 2008 wealth, 2007 NDVI, 2007 LST, and 2007 and 2009 armed conflicts. Density is parameterized such that rate ratios correspond to a 50% increase in density. RR: rate ratio; CI: credible interval, taken over 1,000 draws from the posterior distribution of each parameter; Naive: model fit without accounting for measurement error in livestock density or wealth; MEC: measurement error model.

## Discussion

In Malawi, Uganda, and DRC, we did not find evidence of an indirect effect for NDVI or LST. In South Sudan we did detect a strong indirect effect for LST for both cattle and pigs, and for NDVI for pigs. Direct effects were positive for cattle and pigs in rHAT foci in Uganda and for pigs in gHAT foci in Uganda and DRC, indicating a reservoir effect. In gHAT foci in South Sudan, the direct effect of pigs not mediated by NDVI was positive, but the direct effect not mediated by LST was negative, providing mixed evidence of a reservoir effect for pigs. Direct effects were negative for cattle and pigs in rHAT foci in Malawi, and for cattle in gHAT foci in Uganda and South Sudan. In DRC, direct effects for cattle in gHAT foci in DRC were positive.

These results indicate the environmental pathway mediated by NDVI and LST is weak—if present—in rHAT and gHAT foci in Malawi, Uganda, and DRC. While point estimates were strong in South Sudan, uncertainty was generally large in this country, and these findings may furthermore be due to ecological bias (discussed in detail below). For rHAT, our results suggest livestock are an important reservoir in Uganda but not Malawi, where the zooprophylactic effect predominates. Our findings in Malawi are not necessarily discordant with what is known mechanistically—i.e., cattle and pigs are rHAT reservoirs—if this reservoir poses little risk to humans, allowing the reservoir effect to be dominated by the zooprophylactic effect. This could arise if wildlife are the dominant source of human infection—due to host abundance, tsetse fly preference, contact patterns between hosts, adequate control of the domestic reservoir through insecticides or trypanocides, or other means—and domestic animals exert an important zooprophylactic effect. The spatial distribution of HAT in Malawi, where most active transmission occurs near protected areas, indicates this scenario may be consistent with HAT epidemiology in this setting [43]. For gHAT, our results point to a reservoir role for pigs in both Uganda and DRC and provide some evidence that pigs are a reservoir in South Sudan. These results also provide equivocal evidence that cattle may serve as a gHAT reservoir: while there is evidence of a slight reservoir effect for gHAT in DRC, in Uganda and South Sudan a zooprophylactic effect was detected.

As with any imperfect study—interventional or observational, randomized or not—our results may also be the result of bias. Unmeasured confounding due to omission of vector control from our models, incomplete control by wealth as our parameterization might not have captured the dimension most relevant to our research question, and the need to drop several confounders from our Malawi model due to overfitting concerns, may lead to residual confounding.

Selection bias may arise if clusters or counties excluded from the study are actually at risk of reporting a HAT case, and differ systematically in their distribution of the exposure (livestock density), mediators (LST and NDVI) or confounders (index events, elevation, wealth, vector control). In South Sudan, this could arise if underreporting in the outcome data (discussed below) results in exclusion of an at-risk county from our analyses completely.

One of the major sources of bias in our study is underreporting in the outcome data. If such underreporting is non-differential with respect to HAT risk (i.e., clusters or counties counties at high risk of HAT are no more likely to underreport than those at low risk, even if high-risk individuals are less likely to enter the reporting system than low-risk individuals), effect estimates will be biased to the null in expectation. While weak surveillance infrastructure may drive an association between cluster- or county-level HAT risk and HAT reporting, we expect this mechanism to largely arise via civil unrest, which we have attempted to control for.

Furthermore, with the exception of South Sudan, we have not addressed uncertainty in livestock density and wealth in our estimates, and in South Sudan our implementation of the measurement error model assumes errors in wealth and livestock density are not correlated in space, which is likely violated. Measurement error may also arise in our confounders, driving residual confounding, and in our mediators (NDVI and LST), biasing direct and indirect effects.

Finally, ecological bias arises from aggregation of variables, the presence of an unmeasured individual-level confounder whose association with exposure differs across groups, or the presence of an unmeasured individual-level effect modifier whose effect or distribution differs across groups [44–46]. As ecological bias is sensitive to grouping definition, we are particularly concerned about its effects in South Sudan, where spatial units were large and irregular.

## Conclusion

Concerns about overfitting and residual confounding in the Malawi analyses, ecological bias in the South Sudan analyses, and underreporting in the outcome data for all countries aside, our study takes a novel approach to estimating the pathway-specific effects by which cattle and pigs come to bear on gHAT and rHAT risk. By conducting our study across four high-burden countries, our findings lend strong supporting evidence in favor of a reservoir effect of pigs for gHAT. Our results also indicate that interventions on domestic animal reservoirs have the potential to improve rHAT control in Uganda but are unlikely to yield much benefit in Malawi, where efforts should instead be targeted to vector control and strengthening passive surveillance systems. Control measures targeting the environmental change pathway are unlikely to be of much utility in these countries, their logistical and ethical challenges notwithstanding.

In conjunction with high-resolution livestock maps (S1 File) these results can be interpreted on a policy-relevant scale, allowing policymakers and other stakeholders to identify priority areas for implementing domestic animal AAT control in a coordinated One Health framework.

## Data Availability

All relevant data are within the manuscript and its Supporting Information files

## Supporting information

**S1 Appendix. Wealth mapping**. Methods and results

**S2 Appendix. Motivation and implementation of the mediational g-formula**.

**S3 Appendix. Implementation of the measurement error model in South Sudan**.

**S4 Appendix. Descriptive statistics figures**.

**S1 File. Livestock mapping manuscript**. PDF of author manuscript containing detail on methodology for livestock mapping.

## Acknowledgments

The authors wish to express sincere appreciation to the World Health Organization’s Department of Control of Neglected Tropical Diseases for use of their Atlas of HAT data, and to the HAT research community, in particular Prof. Eric Fevr’e and Dr. Richard Selby.

