## Supplementary material for "Livestock, pathogens, vectors, and their environment: a causal inference-based approach to estimating the pathway-specific effect of livestock on human African trypanosomiasis risk": S1_Appendix

### S1 Appendix: Wealth mapping

##### *Data collection and processing*

Data sources used for livestock and wealth mapping are presented in Table 1.1. There were 10,330 clusters in the final dataset for Malawi, 4,361 in Uganda, and 1,164 in DRC. After reading in data, we harmonized variables so that the wealth index could be calculated across surveys (as detailed in the following section), allowing for smoothing in time.

Table 1.1: Data sources by country

| Country | Year | Source |
|---|---|---|
| Malawi | 2000 | Demographic and Health Survey |
| | 2003 | World Health Survey |
| | 2004 | Second Integrated Household Survey |
| | 2004-2005 | Demographic and Health Survey |
| | 2006 | Technology Adoption and Risk Initiative Survey |
| | 2010 | Demographic and Health Survey |
| | 2010-2011 | Third Integrated Household Survey |
| | 2013, 2016 | Integrated Household Panel Survey |
| | 2012 | Malaria Indicator Survey |
| | 2013-2014 | Multiple Indicator Cluster Survey |
| | 2014 | Malaria Indicator Survey |
| | 2015-2016 | Demographic and Health Survey |
| | 2016-2017 | Fourth Integrated Household Survey |
| | 2017 | Malaria Indicator Survey |
| South Sudan | 2008 | Population and Housing Census |
| DRC | 2007 | Demographic and Health Survey |
| | 2010 | Multiple Indicator Cluster Survey |
| | 2013-2014 | Demographic and Health Survey |

Table 1.1: Data sources by country

| Country | Year | Source |
|---|---|---|
| Uganda | 2001 | Demographic and Health Survey |
| | 2006 | Demographic and Health Survey |
| | 2009 | Malaria Indicator Survey |
| | 2009-2010 | National Panel Survey |
| | 2010-2011 | National Panel Survey |
| | 2011 | AIDS Indicator Survey |
| | 2011 | Demographic and Health Survey |
| | 2011-2012 | National Panel Survey |
| | 2014-2015 | Malaria Indicator Survey |
| | 2016 | Demographic and Health Survey |
| | 2018 | Malaria Indicator Survey |

***Generating the wealth index***

Our process for wealth mapping followed that laid out in the DHS Wealth Index [1] handbook, with the exception of exclusion of livestock-related variables in our wealth index. The final variables included in our wealth models were, defined at the level of the household (M: Malawi, U: Uganda, S: South Sudan, D: DRC):

- Presence of a domestic servant *(M, U, D)*
- Ownership of agricultural land *(M, U, S, D)*
- Amount of land owned (converted to acres) *(M, U, S, D)*
- Ownership of dwelling *(M, U, S, D)*
- Type of fuel used for cooking *(M, U, S, D)*
- Water source *(M, U, S, D)*
- Toilet type *(M, U, S, D)*
- Whether toilet is shared with other households *(M, U, D)*

- Main floor material of dwelling *(M, U, D)*
- Main material of dwelling walls *(M, U, D)*
- Roof material *(M, U, D)*
- Presence of electricity *(M, U, D)*
- One or more household members has a bank account*(M, U, D)*
- Vehicle (none; animal cart or bicycle; motorcycle or scooter; car, mini-bus or truck/lorry) *(M, U, D)*
- Number of rooms per household member *(M, U, S, D)*
- Household owns one or more:
  - Chairs *(M, U, D)*
  - Table *(M, U)*
  - Clock or watch *(M, U, D)*
  - Bucket *(M)*
  - Clothes washing machine *(M)*
  - Dish washing machine *(M, U, D)*
  - Refrigerator *(M, U, S, D)*
  - Mobile phone *(M, U, S, D)*
  - Landline (non-mobile phone) *(M, U, S, D)*
  - Television *(M, U, S, D)*
  - Computer *(M, U, S, D)*
  - Radio *(M, U, S, D)*
  - Sewing machine *(M, D)*
  - Bed *(M, U, D)*
  - Upholstered couch, sofa, chairs, or set *(M, U)*
  - Paraffin or kerosene lamp *(M, U, D)*
  - Boat *(M, U, S, D)*
  - Fishing net *(M)*
  - Mortar and pestle *(M)*
  - Fan *(M, S*
  - Air conditioning *(M)*
  - Machine to play cassette tapes, CDs, DVDs, or Hi-Fi *(M, U)*

  - VCR *(Ms)*
  - Kerosene or paraffin stove *(M)*
  - Electric or gas stove/hot plate *(M, D)*
  - Beer-brewing drum *(M)*
  - Coffee table for sittingroom *(M)*
  - Cupboards, drawers, bureau *(M, U)*
  - Desk *(M)*
  - Iron for pressing clothes *(M)*
  - Satellite dish *(M, S)*
  - Solar panel *(M, U)*
  - Generator *(M, U, D)*
  - Plough *(M)*
  - Tractor *(S)*
  - Axe or hoe *(D)*

- Proportion of possessions enumerated by the survey in question that the household owns *(M, U, D)*

We coded all variables such that a lower level corresponded to lower wealth, and a higher lever corresponded to greater wealth (e.g., we used the variable number of rooms per household member rather than number of household members per room, as the latter would have a negative association with wealth). We assigned levels to categorical variables as detailed in Tables 1.2-1.7; note apparent repetition within variable levels reflect different categorizations across surveys.

**Factor analysis** For Malawi, Uganda, and DRC, we performed exploratory factor analysis separately for each of three periods, to reflect the expectation that factor loadings would change over time (e.g., in early periods ownership of a mobile phone would be much more strongly associated with wealth than in later periods): 2000-2006, 2006-2012, and 2012-2020. For South Sudan, data were only available at the county-level and for a single year (2008), thus wealth mapping was conducted at the county-level for 2008.

First, we removed all variables with more than 5% missingness, and all variables with standard deviation equal to 0. We then created a correlation matrix using the `mixedCor`

Table 1.2: Levels for cooking fuel

| Level | Fuel types |
|---|---|
| | Malawi, Uganda., DRC |
| 4 | Electricity |
| 3 | Gas, kerosene, liquefied petroleum gas, biogas, natural gas, paraffin, oil, solar |
| 2 | Charcoal, coal, lignite |
| 1 | Wood, firewood (collected or purchased) |
| 0 | Dung, straw, shrub, grass, crop residue, saw dust |
| | South Sudan |
| 4 | Electricity |
| 3 | Gas |
| 2 | Charcoal |
| 1 | Firewood |
| 0 | Cow dung, Grass |

Table 1.3: Levels for water source

| Malawi, Uganda, DRC | |
|---|---|
| Level | Water source |
| 4 | Piped into own dwelling, compound, yard or plot |
| 3 | Public or communal standpipe/tap, piped into neighbor's dwelling |
| 2 | Private (in own dwelling, yard or plot) or public protected well, tube well, borehole, protected spring, rain tank or rainwater, channeled by gravity flow scheme |
| 1 | Tanker truck/bowser, cart with small tank, bottled or sachet water, water vendor |
| 0 | Unprotected well or spring, private (in own dwelling, yard or plot) or public open well, river, dam, lake, pond, stream, canal, irrigation channel |
| South Sudan | |
| 3 | Water filtering stations with common standpipe, mechanical boreholes with common standpipe, sand filter with common standpipe, |
| 2 | Deep borehole with network, deep borehole without network, hand pump, water vendor from deep borehole |
| 1 | Shallow well, hafeer/dam with filter, water vendor from shallow well/pond/river/spring |
| 0 | Hafeer/dam without filter, still (turda/fula/river) or running (river/pond/tura'a) open water source |

Table 1.4: Levels for toilet type

| Level | Toilet type |
| --- | --- |
| | Malawi, Uganda, DRC |
| 3 | Flush toilet (piped, pour flush, or unspecified) |
| 2 | VIP latrine, composting toilet, Ecosan toilet, latrine with slab and/or roof |
| 1 | Latrine without slab or roof, public pit toilet or latrine, bucket, hanging toilet |
| 0 | No facilities/open, bush, field |
| | South Sudan |
| 5 | Private flush toilet |
| 4 | Shared flush toilet |
| 3 | Private pit latrine |
| 2 | Shared pit latrine |
| 1 | Bucket |
| 0 | No facilities |

Table 1.5: Levels for dwelling floor

| Level | Floor material |
| --- | --- |
| | Malawi, Uganda, DRC |
| 2 | Carpet, parquet/polished wood, ceramic or other tile or mosaic, cement/concrete, smooth cement, vinyl/asphalt strips, linoleum, stone, bricks |
| 1 | Palm/bamboo, wood planks, broken bricks |
| 0 | Earth, sand, dung, smoothed mud |

Table 1.6: Levels for dwelling walls

| Level | Wall material |
|---|---|
| | Malawi, Uganda, DRC |
| 3 | Cement, concrete, stone with lime/cement or mud, finished/burnt brick with stone or mud, covered adobe |
| 2 | Wood, wood planks/shingles, reused wood, plywood, plastic sheet, metal sheet, corrugated iron |
| 1 | Mudbrick, unburnt bricks with plaster, cement, or mud, uncovered adobe |
| 0 | Thatch, straw, grass, cane/palm/trunks, bamboo or poles with mud, timber, mud, dirt, compacted earth, cardboard, none |

Table 1.7: Levels for dwelling roof

| Level | Roof material |
|---|---|
| | Malawi, Uganda, DRC |
| 3 | Roofing shingles, ceramic tiles, clay tiles, cement, concrete, calamine/cement fiber, wood, asbsetos |
| 2 | Iron sheets, metal, tin |
| 1 | Wood planks, cardboard, palm/bamboo grass, rustic mat, plastic/polythene sheeting, tin cans |
| 0 | Thatch/palm leaf, grass, sod, mud/earth, none |

function in the `psych` package in R [2], and performed exploratory factor analysis using the `fa()` function in the same package, setting our arguments to those specified by the DHS Wealth Index manual: principal components extraction with one factor extracted, imputation of mean for missing data, and estimation of the factor scores using the regression method.

We performed linear regression of the extracted scores on input variables as a sanity check. For all four countries, in general only variables with very large levels of missingness had negative associations with our final wealth score. In all countries, ownership of land was negatively associated with our final wealth score and had high levels of missingness (17% missing in Malawi, 25% in South Sudan, 15% in Uganda, and 31% in DRC). Similar results were found for ownership of dwelling (negative association in Malawi with 87% missing, South Sudan with 0% missing, DRC with 93% missing), area of land owned (negative association in South Sudan with 25% missing, Uganda with 92% missing, and DRC with 93% missing), and household sharing toilet with other households (negative association in Uganda with 12% missing, DRC with 15% missing). In DRC, negative associations were also found for ownership of a canoe, axe, or hoe, with 0% missing for these variables, reflecting absence of a strong association between ownership of these items and wealth. All other variables had positive associations with the final wealth score.

As the goal was to adjust for wealth score in final regression models, we did not perform factor analysis separately for urban and rural clusters in Malawi, Uganda, or DRC, however we use urban/rural status as a predictor in our models (discussed in the Mapping section, below) to reflect both the expected association between this variable and wealth score, and the stratified design of the surveys we used.

##### ***Mapping***

We detail our approach to livestock mapping in Malawi, Uganda, DRC, and South Sudan using these data in an accompanying publication, available as a preprint in S1 File. This file contains details on the stochastic partial differential equations (SPDE) to Gaussian process modeling adopted in countries with point-level data (Malawi, Uganda, and DRC), and the small area estimation approach adopted in South Sudan.

**Malawi, Uganda and DRC** After the wealth index was constructed, we collapsed over cluster by taking the mean wealth score in each cluster.

Our predictors included urban/rural status to reflect sampling strategy, and nighttime lights. Data on nighttime lights came from the National Oceanographic and Atmospheric Administration's Nighttime Lights Time Series, which are annual cloud-free composites made from archived DMSP-OLS data, available at a resolution of 30-arc-seconds [3]. These data are available from 1992-2013, as average visible lights, stable lights, and a normalized version of average lights. We used average visible lights, and for model fitting and prediction for 2014-2020 we used data from 2013.

In addition to model selection via leave one out cross-validation as detailed in S1 File, for external validation we also performed spatial regression to check for association between our final wealth index and the proportion of the population earning under $2 per day in 2010. These latter estimates are produced by WorldPop using Bayesian model-based geostatistics and household survey data from the LSMS program, and are available at a resolution of $0.00833^{\circ}$ [4]. We found that a one unit increase in proportion earning less than $2/day (i.e., going from 0 to 1) was associated with a 0.84 lower wealth score as predicted by our final Malawi model and a 0.87 lower wealth score as predicted by our final Uganda model, indicating good agreement between our wealth maps and the WorldPop poverty maps. WorldPop does not produce these estimates for DRC.

**South Sudan** In South Sudan we produced direct estimates of county-level wealth and design-based variance using the `svydesign()` and `svyby()` functions in the `survey` package and household weights provided in the IPUMS extract. In contrast with our livestock mapping methodology dtailed in S1 File we did not perform smoothing as our primary goal was not to estimate wealth, thus stabilizing variance of this estimate was not critical.
