## Supplementary material for "Livestock, pathogens, vectors, and their environment: a causal inference-based approach to estimating the pathway-specific effect of livestock on human African trypanosomiasis risk": S2_Appendix

### S2 Appendix: Motivation and implementation of the mediational g-formula

##### *Identifiability criteria*

By notational convention, let $L$ be a vector of measured confounders, $A$ be exposure, $Y$ be outcome, $M$ be mediator(s), and $T$ be time, and let overbars denote history through time $T = t$. The identifiability criteria are exchangeability, positivity, and the stable unit treatment value assumption (SUTVA).

Exchangeability refers to the absence of confounding. As this is rarely satisfied marginally, the weaker assumption of exchangeability conditional on the past and on $L$ is typically adopted. Using counterfactual notation: (1) no exposure-outcome confounding, conditional on the past: $Y^{\overline{a}M^{\overline{a}}} \perp\!\!\!\perp A_t|\overline{A}_{t-1}, \overline{L}_t$,(2) no mediator-outcome-confounding, conditional on the past: $Y^{\overline{a}M^{\overline{a}}} \perp\!\!\!\perp M_t|\overline{A}_t, \overline{M}_{t-1}, \overline{L}_t$, and (3) no exposure-mediator confounding, conditional on the past: $M^{\overline{a}} \perp\!\!\!\perp A_t|\overline{A}_{t-1}, \overline{M}_{t-1}, \overline{L}_t$ [1].

Positivity refers to a positive probability of being assigned to each of the treatment levels and is expressed as $Pr(A = a|L = l) > 0$ for $Pr(L = l) \neq 0$.

SUTVA has two components: consistency and no interference. Consistency refers to a sufficiently well-defined exposure; that is, each unit of observation (here, a cluster-year) has one potential outcome for a given treatment level $A = a$, implying a cluster-years observed outcome equals their counterfactual outcome under their observed exposure: if $A_i = a$, then $Y_i^a = y_i$ for all clusters. Under no interference, each cluster's potential outcome is independent of all other clusters' potential outcomes. In our study we instead assume partial interference, detailed in the main text.

##### *Implementation*

The causal estimands are defined in the main text. We implemented all models using the `glm()` function in R; we did not use spatial models due to computational challenges and concerns regarding spatial confounding. We fit Gaussian models for wealth, NDVI, and

LST, and Poisson models for HAT cases. Our procedure was as follows, iterating over each $t$ with $t_0$ defined as 2001 (2000 was not modeled due to the need to implement lags):

1. For $t_0$, fit all models except the outcome model (HAT cases): wealth, NDVI, and LST. Outcome model not fit due to the need to lag livestock by two time points (years)
2. Set livestock density to $A = a$, and for each model in turn use the `predict()` function to predict the variable in question under $A = a$ on the Monte Carlo sample.
   (a) If indicated by the counterfactual parameter in question, instead set $A = a^*$ when predicting from the mediator model
3. Repeat steps 1-2 for all times $t_1, ..., t_T$, adding in the outcome model for $t > 0$.
4. Iterate steps 1-3 over each bootstrapped sample

The result of this procedure is a vector of counterfactual outcomes for each simulation $s$, with $Y_{t(s)i}^{\overline{a}M^{\overline{a}}}$ defined as the 1-year cumulative incidence of HAT in cluster $i$, year $t$, simulation $s$, and with exposure set to $\overline{A} = \overline{a}$.

$$\mathbf{Y}_{(s)i}^{\overline{a}M^{\overline{a}}} = \left\{ Y_{1(s)i}^{\overline{a}M^{\overline{a}}}, Y_{2(s)i}^{\overline{a}M^{\overline{a}}}, ..., Y_{T(s)i}^{\overline{a}M^{\overline{a}}} \right\}$$

As noted above, we contrast counterfactual outcomes on the ratio scale. Taking the natural indirect effect as an example, across clusters, time and simulations, we implement this as follows, separately for each country (and for gHAT and rHAT in Uganda):

$$E_{(s)}\left[ E_t\left[ E_i[Y_{t(s)i}^{\overline{a}M^{\overline{a}}}] \,/\, E_i[Y_{t(s)i}^{\overline{a}M^{\overline{a}^*}}] \right]\right]$$

with uncertainty bounds estimated by taking the 2.5% and 97.5% quantiles across simulations.
