## Supplementary material for "Livestock, pathogens, vectors, and their environment: a causal inference-based approach to estimating the pathway-specific effect of livestock on human African trypanosomiasis risk": S3_Appendix

### S3 Appendix: Implementation of the measurement error model in South Sudan

The measurement error (MEC) model is defined as:

$$w_c = x_c + u_c, c = 1, ..., n$$

where $c$ indexes county, $u_c$ are the measurement error terms distributed as $u_c \sim N(0, \sigma_u^2)$, and $x_c$ and $u_c$ are independent. Since the $u$ terms have mean 0, $E(W|X = x) = x$, that is, $W$ is unbiased for a given unobserved $x$. Failing to account for measurement error will result in biased effect estimates and inappropriate standard errors [1].

Following the heteroscedastic errors-in-variables approach detailed in Wang et al. (2018), we defined our hierarchical model as follows:

$$\lambda = exp\left(\theta_0 + \theta_1\mu_c + \theta_2 m_c + \theta_3\eta_c + \boldsymbol{\alpha}\mathbf{z}(\mathbf{k}_c) + S_c + \epsilon_c + log(P_c)\right)$$

$$\mu_c = x_{lc} + u_{lc}, \eta_c = x_{wc} + u_{wc}$$

$$x_{lc} = \lambda_l + \zeta_{lc}, x_{wc} = \lambda_w + \zeta_{wc}$$

$$u_{lc} \sim N(0, d_c\sigma_{ul}^2), u_{wc} \sim N(0, d_c\sigma_{uw}^2)$$

$$\zeta_{lc} \sim N(0, \sigma_{xl}^2), \zeta_{wc} \sim N(0, \sigma_{xw}^2)$$

$$S_c|S_k, k \in ne(c) \sim N\left(\overline{S}_k, \frac{\sigma_s^2}{n_c}\right)$$

$$\epsilon_c|\sigma_\epsilon^2 \sim_{iid} N(0, \sigma_\epsilon^2)$$

where

- $\mu_c$ is estimated livestock density in county $c$ in 2008
- $m_c$ is the mediator value in county $c$ in 2010, $M \in$\{NDVI, LST\}
- $\eta_c$ is estimated wealth score in county $c$ in 2008
- $\boldsymbol{\alpha}$ is a vector of coefficients

- $\mathbf{z}(\mathbf{k}_c)$ is a vector of exposure-outcome, exposure-mediator, and mediator-outcome confounders measured without error, $k \in$ {2007 NDVI, LST, conflicts, disasters; 2009 conflicts, disasters}
- $P_c$ is the offset, given as population in county $c$
- $S_c$ are county-level structured random effects which follow the ICAR model with marginal variance $\sigma_s^2$
- $ne(c)$ denotes neighbors (shared boundary) of county $c$
- $n_c$ is the number of neighbors of county $c$
- $\epsilon_c$ are county-level iid (unstructured) random effects with variance $\sigma_\epsilon^2$
- $x_{lc}$ is the true livestock density in county $c$
- $u_{lc}$ is the measurement error for livestock density in county $c$
- $x_{wc}$ is the true wealth score in county $c$
- $u_{wc}$ is the measurement error for wealth score in county $c$
- $\lambda_l$ is the mean of the true livestock density
- $\lambda_w$ is the mean of the true wealth score
- $\zeta_{lc}$ is the residual for livestock density in county $c$
- $\zeta_{wc}$ is the residual for wealth score in county $c$
- $d_c$ is a weight which allows for heteroscedasticity in the error structure

For both livestock density and wealth score, we assume $x_c \sim N(\theta, \sigma_x^2)$, where $\theta$ is the mean of $x_c$ and $\sigma_x^2$ is the variance.

There are therefore four parameters needed for each of the two resulting measurement error models: $\beta_1$ ($\beta_2$ for wealth score), $\log(1 / \sigma_u^2)$, $\lambda$, $\log(1 / \sigma_x^2)$, as well as the scale factor $d_c$. For livestock density, we specified the priors and starting values for these parameters as follows for livestock, substituting $\mu$ for $\eta$ and $\beta_1$ for $\beta_2$ for wealth:

- $\log(1 / \sigma_u^2) \sim \text{logGamma}(10, 10)$, starting value $\log\big(1 / var(\sigma_\mu)\big)$
- $\beta_1 \sim \text{Normal}(\hat{\beta}_1, 1 / \hat{\sigma}^2_{\hat{\beta}_1})$.
- $\lambda$ fixed at mean $E[\mu]$, with a Gaussian prior
- $\log(1 / \sigma_x^2) \sim \text{logGamma}(10, 10)$, starting value $\log(1 / var(\mu))$
- $d_c \sim \text{Unif}(0.5, 1.5)$

where $var(\sigma_\mu)$ is the empirical variance of the posterior standard deviation for livestock density, $\hat{\beta}_1$ is the posterior mean of the coefficient for livestock density from the naive model, $\hat{\sigma}_{\hat{\beta}_1}$ is the posterior standard deviation of the same coefficient from the naive model, and $var(\mu)$ is the empirical variance of the posterior median for livestock density. The logGamma prior specification is equivalent to that used in Wang et al. [1]. The MEC model for wealth was specified equivalently, and in our final models both livestock density and wealth were included in this form.

The MEC mediator model follows as for the outcome model detailed above.
