## Supplementary material for "Livestock, pathogens, vectors, and their environment: a causal inference-based approach to estimating the pathway-specific effect of livestock on human African trypanosomiasis risk": S4_Appendix

### S4 Appendix: Descriptive statistics plots

***Malawi***

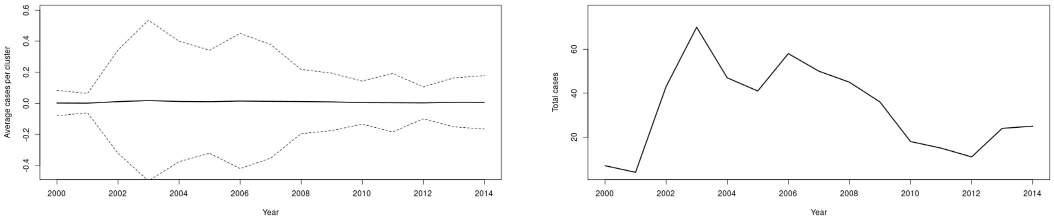


(a) (b)

Figure 4.1: Mean (a) and sum (b) of cases over time in study clusters, Malawi

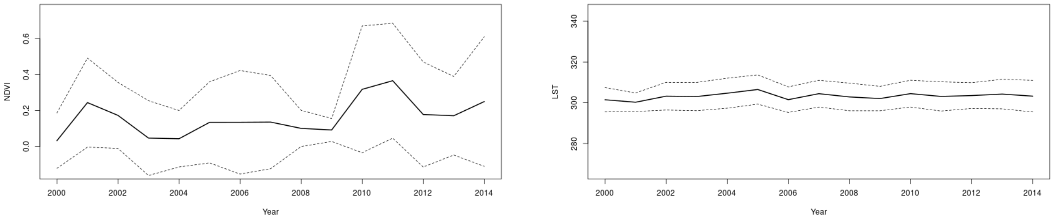


(a) NDVI (b) LST

Figure 4.2: NDVI (a) and LST (b) over time in study clusters, Malawi

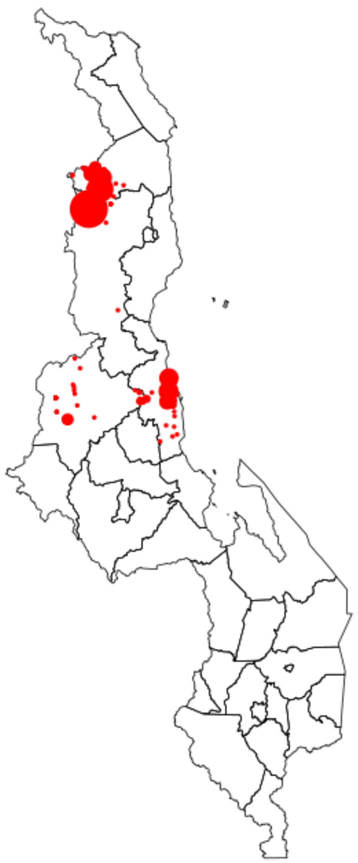

Figure 4.3: HAT cases 2000-2014, Malawi

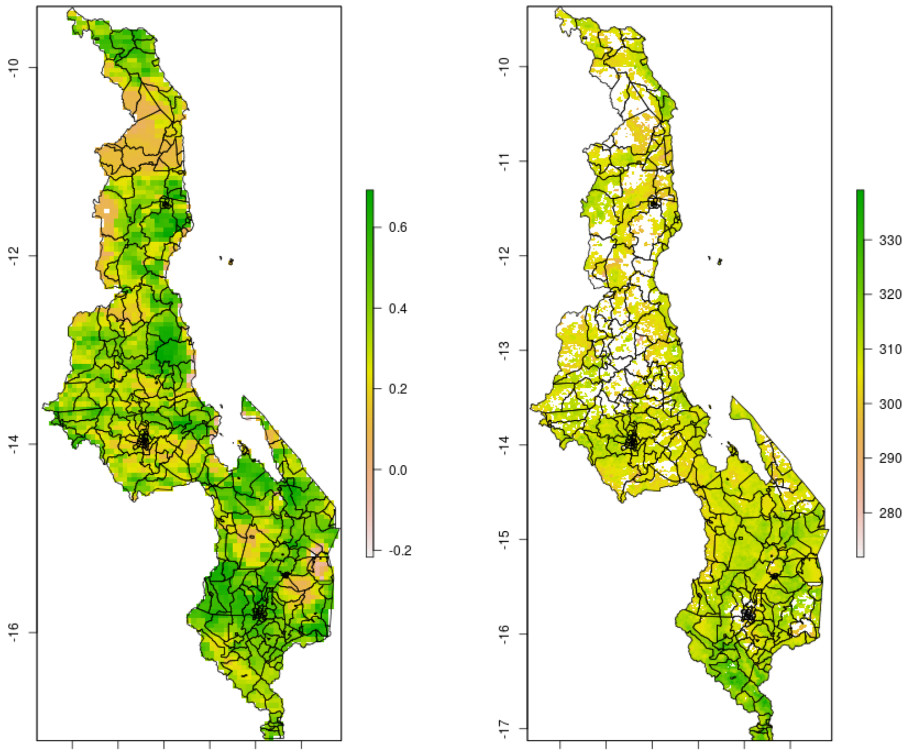


(a) (b)

Figure 4.4: 2010 NDVI (a) and LST (b), Malawi

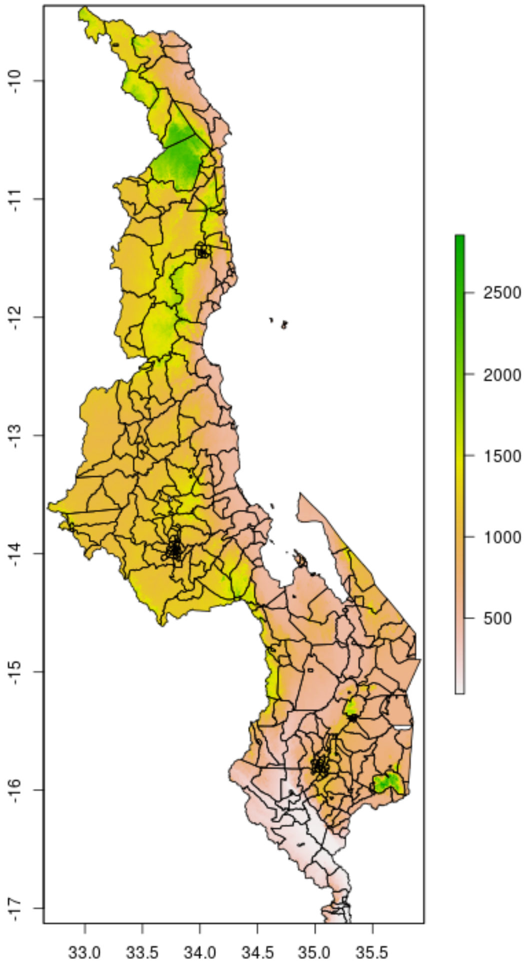


Figure 4.5: Elevation, Malawi

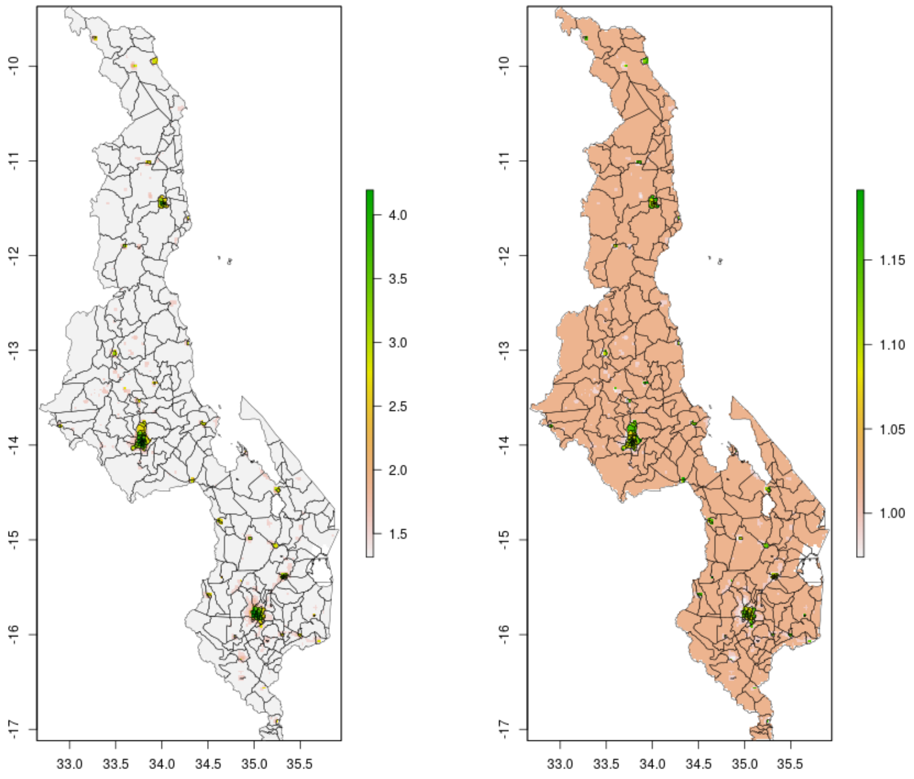


(a) (b)

Figure 4.6: 2010 wealth scores, mean (a) and posterior 95% credible interval (b), Malawi

***Uganda***

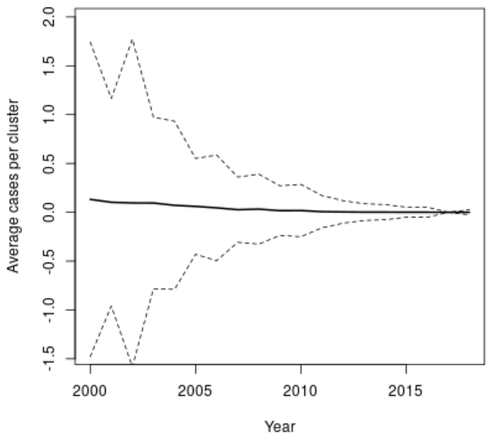


(a) gHAT

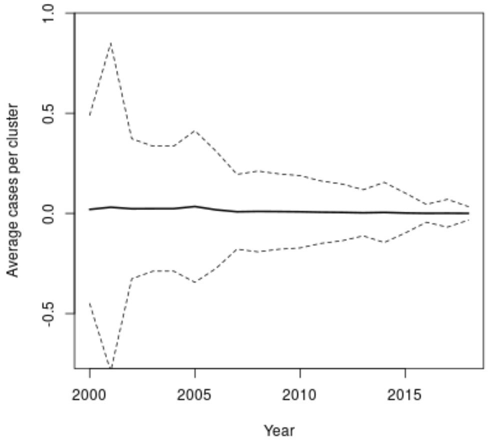


(b) rHAT

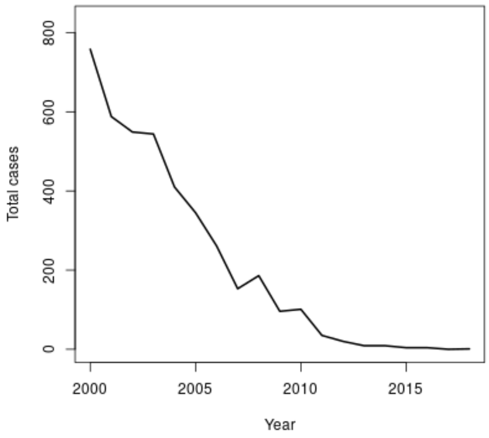


(c) gHAT

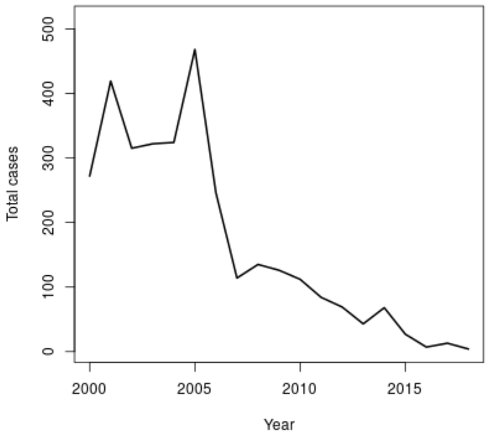


(d) rHAT

Figure 4.7: Mean (a-b) and (c-d) of cases over time in study clusters, Uganda

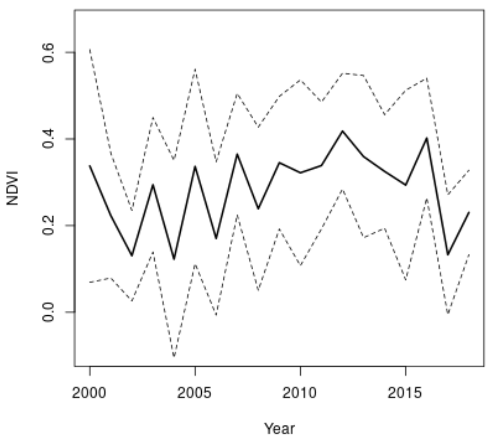


(a) gHAT

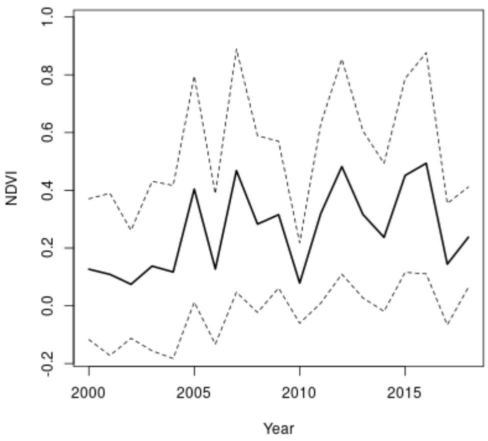


(b) rHAT

Figure 4.8: NDVI over time in study clusters, Uganda

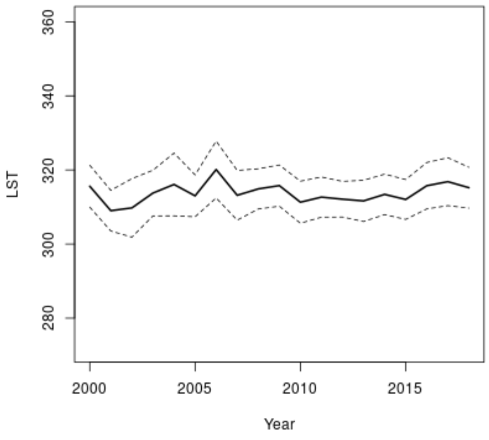


(a) gHAT

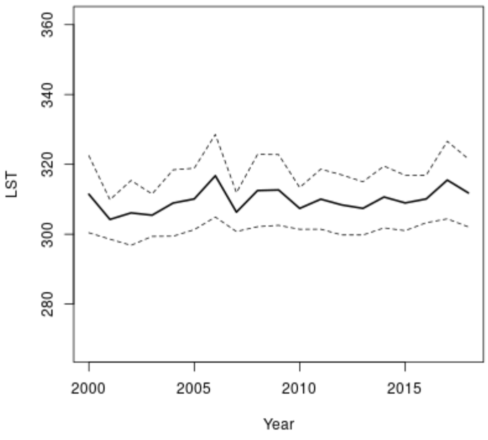


(b) rHAT

Figure 4.9: LST over time in study clusters, Uganda

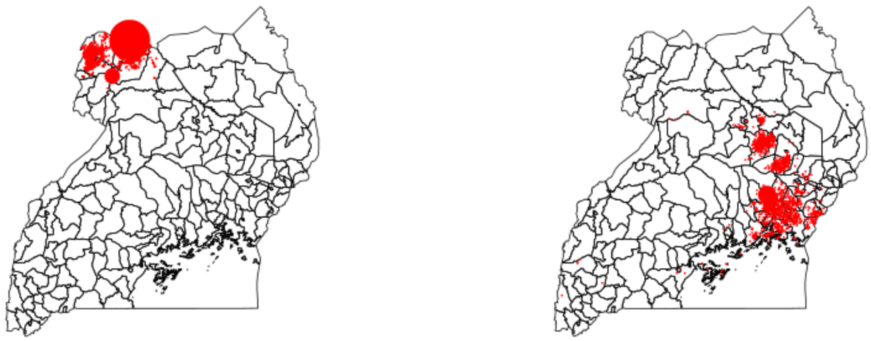

(a) gHAT (b) rHAT

Figure 4.10: HAT cases 2000-2018, Uganda

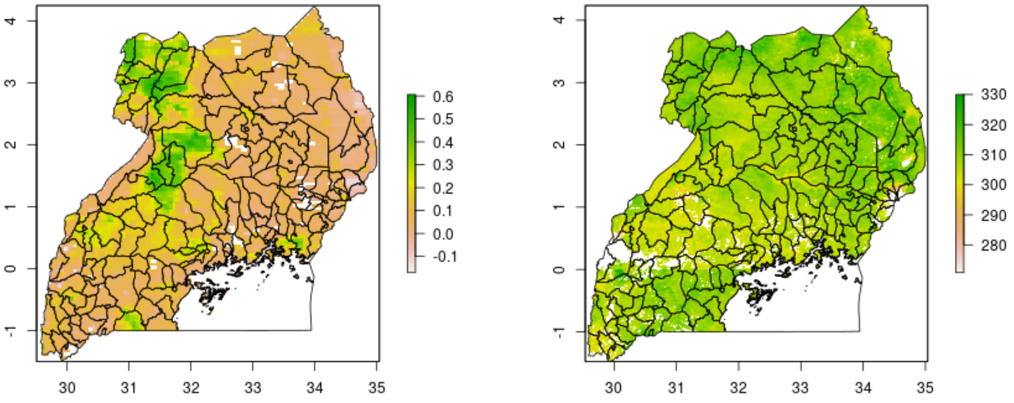


(a) (b)

Figure 4.11: 2010 NDVI (a) and LST (b), Uganda

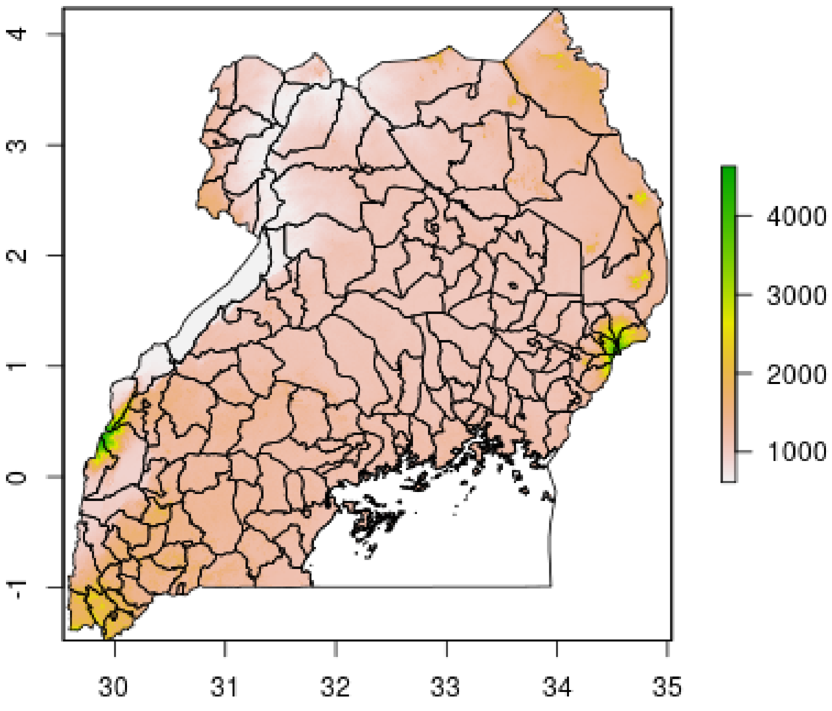


Figure 4.12: Elevation, Uganda

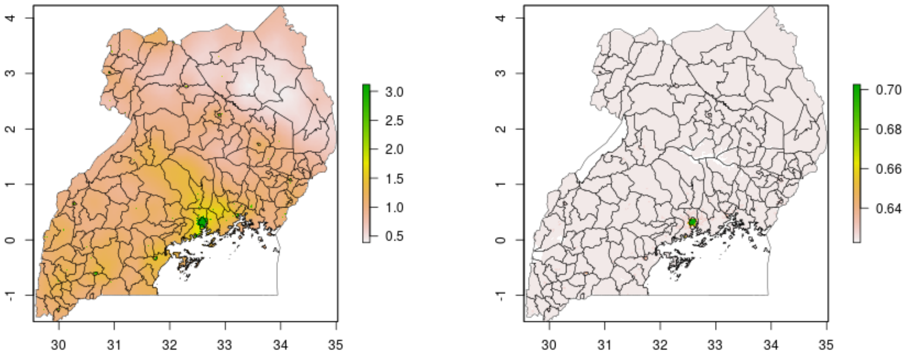


(a) (b)

Figure 4.13: 2010 wealth scores, mean (a) and posterior 95% credible interval (b), Uganda

#### *DRC*

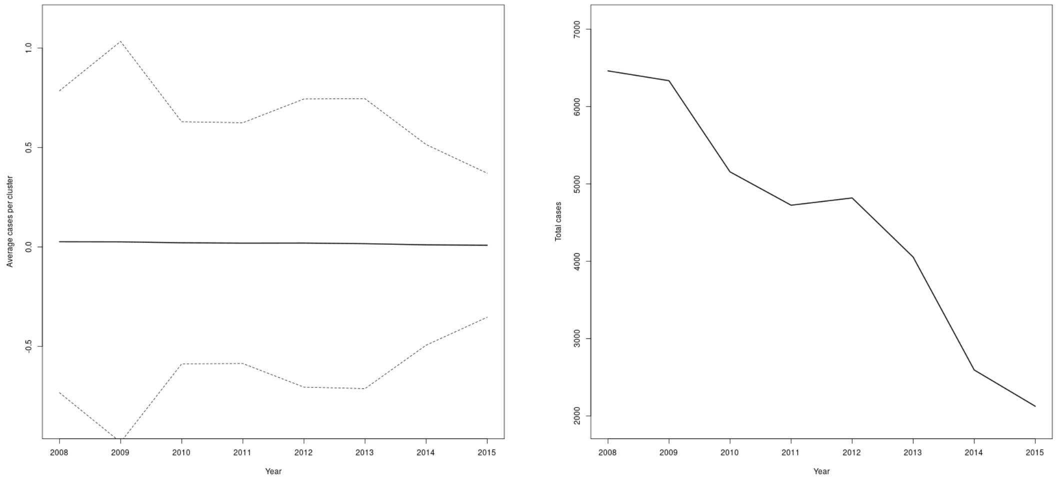


(a) Mean (b) Sum

Figure 4.14: Mean (a) and sum (b) of cases over time in study clusters, DRC

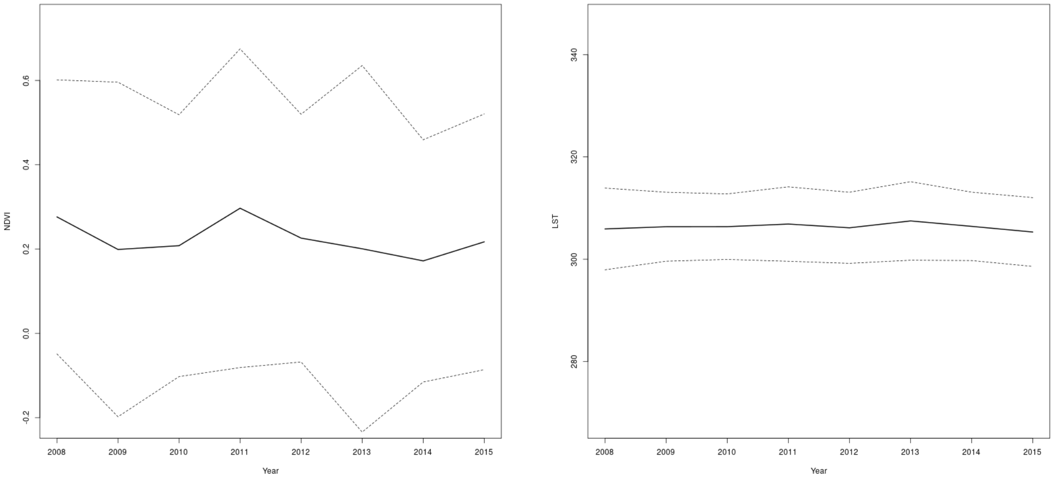


(a) NDVI

(b) LST

Figure 4.15: NDVI (a) and LST (b) over time in study clusters, DRC

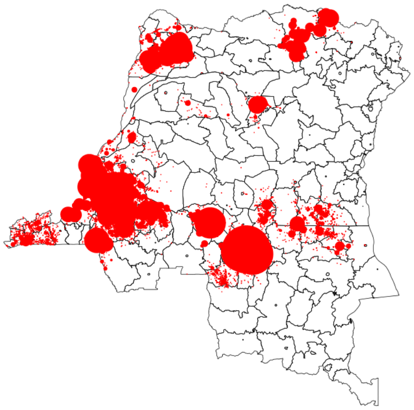

Figure 4.16: HAT cases 2008-2015, DRC

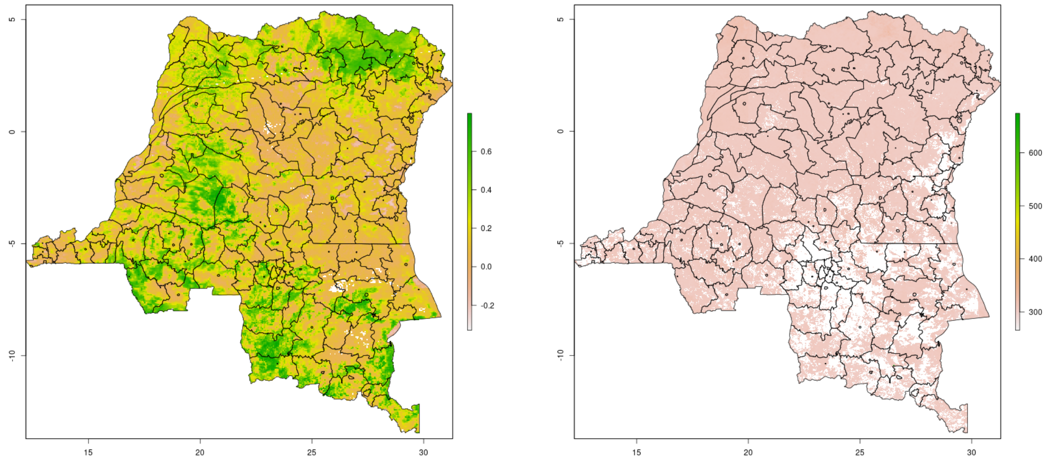


(a) (b)

Figure 4.17: 2010 NDVI (a) and LST (b), DRC

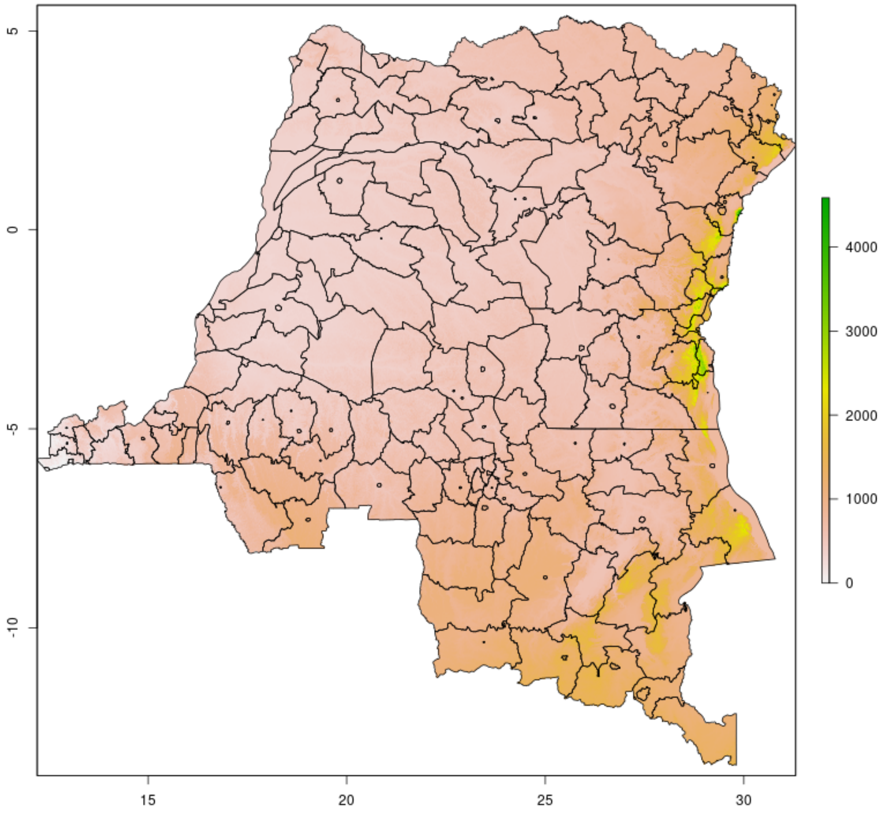


Figure 4.18: Elevation, DRC

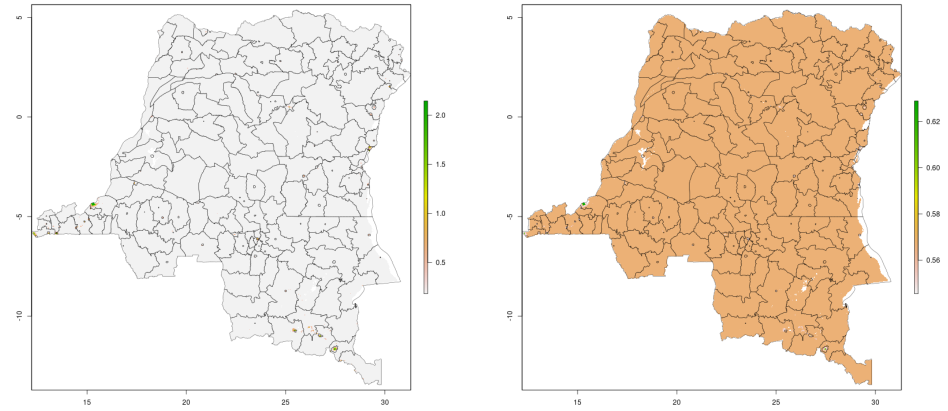


(a) (b)

Figure 4.19: 2010 wealth scores, mean (a) and posterior 95% credible interval (b), DRC

#### ***South Sudan***

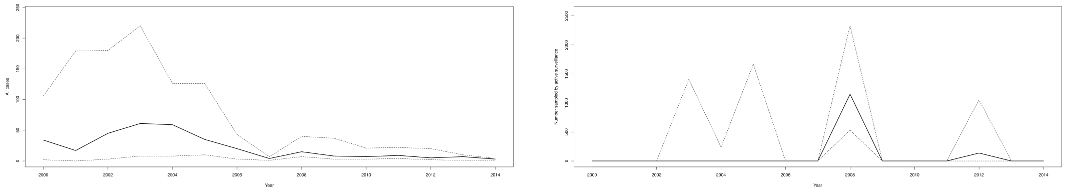


(a) (b)

Figure 4.20: Mean number of cases (a) and people sampled by active surveillance (b) over time in study counties, South Sudan

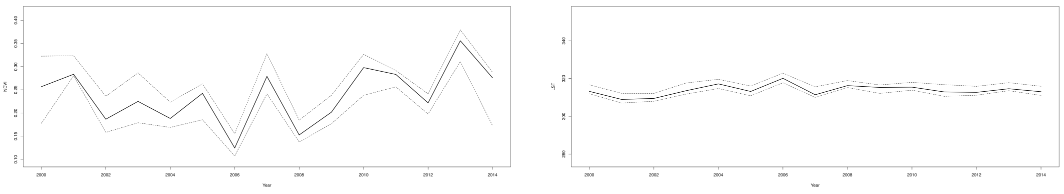

(a) NDVI (b) LST

Figure 4.21: NDVI (a) and LST (b) over time in study counties, South Sudan

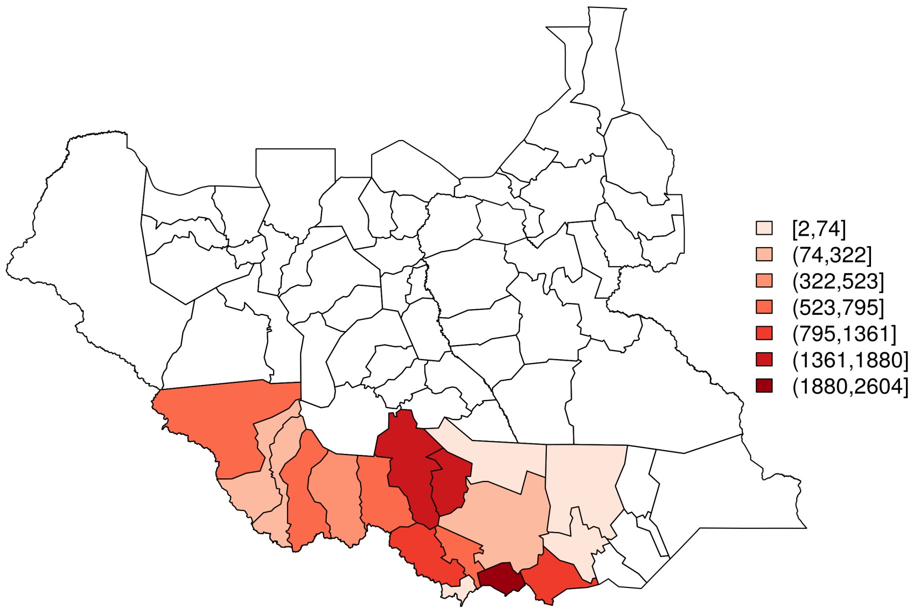


Figure 4.22: HAT cases 2000-2014, South Sudan

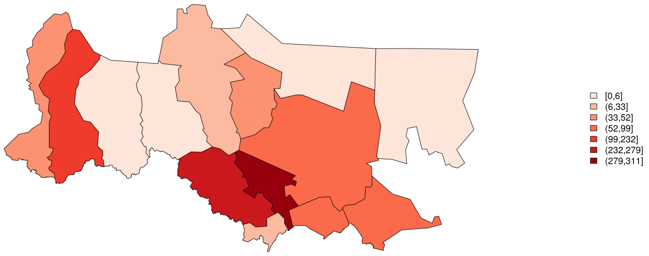


(a) Cases detected

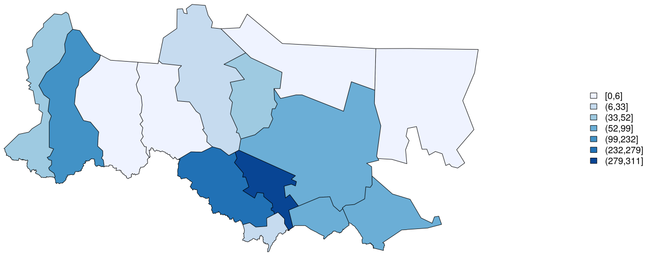


(b) Number sampled

Figure 4.23: Total cases detected (a) and number sampled (b) by active surveillance in South Sudan, 2008 (restricted to active surveillance area)

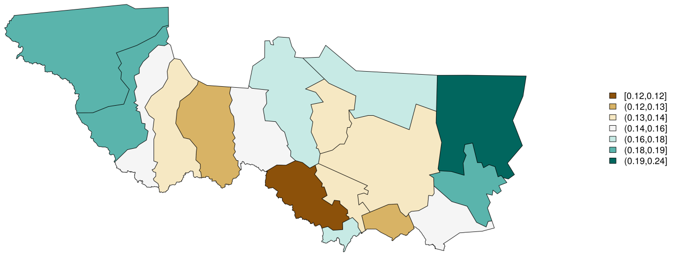


(a) NDVI

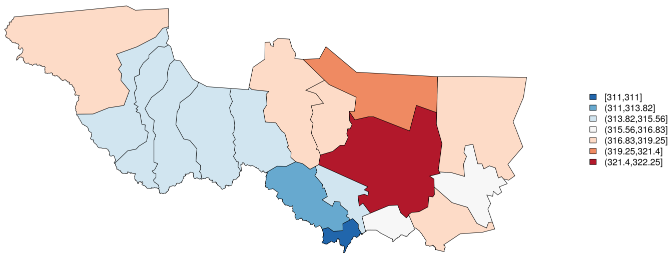


(b) LST

Figure 4.24: 2008 NDVI (a) and LST (b), South Sudan (study area)

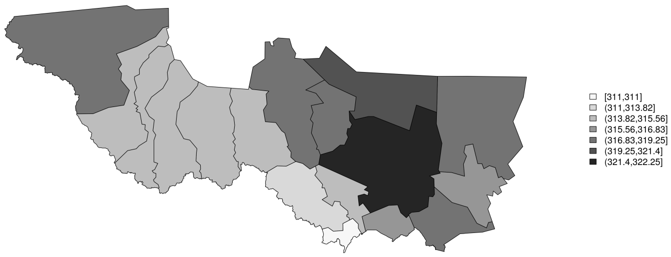


Figure 4.25: Elevation, South Sudan (study counties)

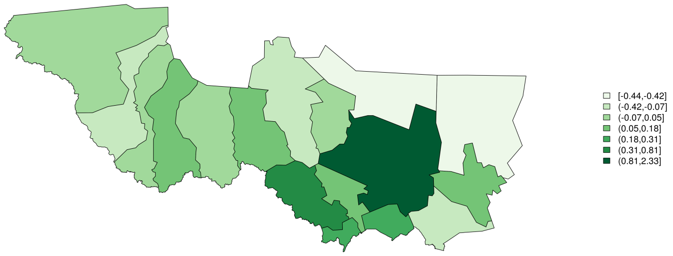


(a) Median

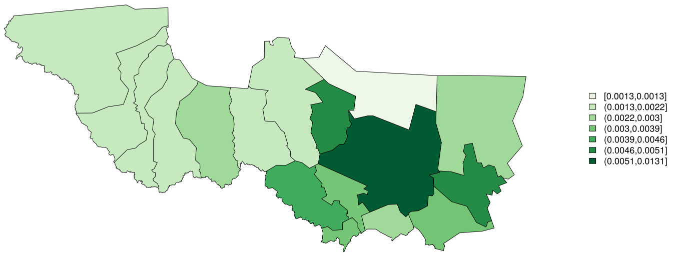


(b) Standard error

Figure 4.26: 2008 wealth, median (a) and standard error (b), South Sudan (study counties)
